# Complex harmonic manifolds in mindfulness-based cognitive therapy for major depressive disorder

**DOI:** 10.64898/2026.06.26.26356643

**Authors:** Paulina Clara Dagnino, Anne Maj van der Velden, Yonatan Sanz Perl, Sara W. Lazar, Henricus G. Ruhe, Jakub Vohryzek, Gustavo Deco, Morten L. Kringelbach

## Abstract

Major depressive disorder (MDD) is a heterogeneous mental disorder characterised by rumination. Mindfulness-based cognitive therapy (MBCT) is an evidence-based treatment developed to target rumination and recurrence risk. Ongoing studies have begun to identify neural changes associated with treatment effects. However, the low-dimensional organisation underlying whole-brain dynamics remains largely unexplored and may provide a more complete characterisation of the neural processes through which MBCT exerts its therapeutic effects in MDD. Here, we investigated functional magnetic resonance imaging (fMRI) of a randomised controlled trial of MBCT with treatment as usual (TAU), or TAU alone, in a group of MDD patients (N=80). We applied a novel framework, complex harmonics decomposition (CHARM), to uncover low-dimensional manifolds in the spacetime domain, capturing local as well as non-local interactions made possible by brain criticality and amplified by the anatomical long-range connectivity. We successfully identified distinct distributed spatiotemporal manifolds across brain states and outperformed traditional dimensionality reduction techniques. During rumination after MBCT we found consistent recruitment of regions involved in bodily and interoceptive processing integrated within the whole-brain across manifolds, changes in latent configurations associated with clinical and behavioural improvements, and greater flexibility within the reduced space. Integration of bodily and interoceptive processing regions within distributed whole-brain manifolds and greater brain flexibility may be associated with reduced “stickiness” of ruminative thinking patterns following mindfulness training in depression. Our findings highlight the promise of low-dimensional manifolds and long-range interactions arising from critical brain dynamics in understanding how mindfulness targets depressive ruminative processing.

**Trial registration:** ClinicalTrials.gov NCT03353493.

## Introduction

Major depressive disorder (MDD) is a leading cause of global disability (WHO, 2017). It is a mental disorder with heterogeneous symptoms characterised by rumination. Mindfulness-based cognitive therapy (MBCT), which combines cognitive behavioural therapy (CBT) and mindfulness practice, is a cost-effective treatment that has demonstrated clinical benefit for recurrent and relapsing depression (Segal, 2013; van der Velden, 2015). A meta-analysis revealed that 62% of individuals were relapse-free over 60 weeks after MBCT delivered either as monotherapy or alongside other treatments, with higher efficacy in individuals with more severe symptoms. Furthermore, it lowered relapse risk by 31% compared with non-MBCT treatments including treatment as usual (TAU), psychological education and antidepressant medication (ADM), and by 23% when compared specifically to ADM (Kuyken et al., 2016). Other studies have shown MBCT reduces relapse risk with equal effectiveness as CBT (Farb et al., 2018; Teasdale et al., 2002). Posterior research has found MBCT can also help during the acute phase of depression (Barnhofer et al., 2025; Goldberg et al., 2019; Pots et al., 2014; Thimm & Johnsen, 2020; Tickell et al., 2020; van Aalderen et al., 2015), as well as in patients with treatment-resistant depression (Cladder-Micus et al., 2018; Eisendrath et al., 2016).

To optimise the therapy, it is key to understand the psychological processes and brain mechanisms underlying the therapy. Neuroscience research in MBCT for MDD is in its early stages and ongoing studies are investigating the neural reconfigurations associated with treatment response. Brain network dysfunction is considered a core feature of depression (Kaiser et al., 2015) and network reorganisation is believed to underlie symptomatology improvements in MBCT (Gkintoni et al., 2025). The main brain networks related to depressive rumination are the salience network (SAL) and the default mode network (DMN). The SAL, associated with interoceptive awareness and emotion and attention regulation, is suggested to be implicated in depression symptomatology and response to different treatments (Downar et al., 2016; Godlewska et al., 2018; Young et al., 2018). On the other hand, the DMN, associated with internal thoughts, self-referential thinking and mind-wandering (Mason et al., 2007), has become the central advocate in depression given the relation between alterations in DMN activity and connectivity with ruminative thoughts (Hamilton et al., 2015; Hamilton et al., 2011; Zhou et al., 2020) and a central target for depression treatment including mindfulness training (Barnhofer & Wild, 2016).

In our clinical trial of MBCT added to treatment as usual (TAU), and TAU alone, we quantified changes in brain function reconfigurations using functional magnetic resonance imaging (fMRI) scans (ClinicalTrials.gov NCT03353493), with effects primarily observed during rumination mind state following MBCT+TAU. We investigated neurocognitive processes during rumination by implementing a static functional connectivity analysis (van der Velden, 2023), a dynamic data-driven network approach (van der Velden, 2025) and a hierarchical brain organisation study (Dagnino et al., 2026) finding distinct neural changes associated with improvements in clinical and behavioural outcomes. Together these analytical approaches have contributed to a more complete characterisation of MBCT induced neural changes in MDD and may be consistent with a shift away from self-reinforcing and entrenched ruminative patterns towards more differentiated and less automatically coupled cognitive and bodily cycles. However, shifting the focus from uncovering brain networks observed in brain dynamics, to capturing the low-dimensional organisation underlying such brain dynamics may provide a more complete characterisation of the neural processes through which MBCT exerts its therapeutic effects in MDD.

The brain is a complex network with high-dimensional neural activity. Brain activity is organised into distinct patterns of functional coordination across regions, enabling the large-scale neural communication necessary for cognition and hypothesised key for brain function (Sporns et al., 2000). Despite decades of research investigating different approaches to describe the high-dimensional space of whole-brain neuroimaging with lower-dimensional features, brain organisation is not trivial and our understanding is still incomplete. The first investigation began with lesion studies (Mesulam, 1990) and progressed to correlational analysis (Biswal et al., 1995), brain activation (Raichle et al., 2001) and mathematical tools such as principal component analysis (Jolliffe & Cadima, 2016) and independent component analysis (McKeown et al., 1998). Later, group-based clustering revealed the widely used Yeo resting-state networks (RSN) (Yeo et al., 2011), and many other approaches such as autoencoders (Sanz Perl et al., 2020) continue being studied until today. A limitation of existing methods is that they do not capture the distributed non-local and spatiotemporal dependencies arising from long-range functional coordination made possible by criticality, and amplified by the anatomical long-range connectivity, proposed to facilitate the time-critical computations required for survival (Deco et al., 2021; Itoh et al., 2022). Brain criticality is a dynamical regime between order and disorder, thought to support maximal information transmission and optimal processing among neural networks (Deco & Jirsa, 2012; Hesse & Gross, 2014; Massobrio et al., 2015), and proposed clinically relevant to depression and persistent rumination (Zimmern, 2020). To this end, (Deco et al., 2025) developed a novel theoretical framework, named complex harmonics decomposition (CHARM), to uncover low-dimensional manifolds capturing non-locality in critical spacetime brain dynamics. This approach has so far been successfully applied to study wakefulness and sleep (Deco et al., 2025). As such, CHARM is a promising framework for uncovering low-dimensional manifolds arising from spacetime critical brain dynamics in depressive rumination, and how mindfulness training may target such processes on a neural level.

Here, we conducted a secondary analysis to explore changes in low-dimensional brain organisation in patients with MDD following MBCT+TAU and TAU using CHARM (Deco et al., 2025). This approach formulates the manifold reduction problem (Belkin, 2003) using the Schrödinger’s equation to uncover the underlying low-dimensional networks that capture not only local but also interferences and aggregation of information transfer mediated by nonlocal brain dynamics arising from criticality, and amplified by anatomical long-range connections in the spacetime domain. In this way, CHARM extends our previous data-driven brain patterns approach (van der Velden, 2025) and brain hierarchy study (Dagnino et al., 2026), by uncovering low-dimensional networks that overlap in time and enabling the quantification of hierarchical organisation and latent configurations within the reduced space. To our knowledge, this is the first study using CHARM to investigate MBCT related brain changes in MDD, opening new avenues to study reorganisation of low-dimensional networks in neuropsychiatric disorders.

## Materials and Methods

### Study design

The study design consisted of a single-blind, randomised controlled trial of MBCT in addition to TAU, or TAU alone, initially published in (van der Velden, 2023) (**Figure 1A**). It was approved by the Regional Ethics Committee of Central Denmark (ID: 1-10-72-259-16: 66534 and registered in the Danish Data Protection Agency (2016-051-000001) and ClinicalTrials.gov (identifier: NCT03353493). For details on participant recruitment (N=80) please see **Supplementary Information**.

**Figure 1.**
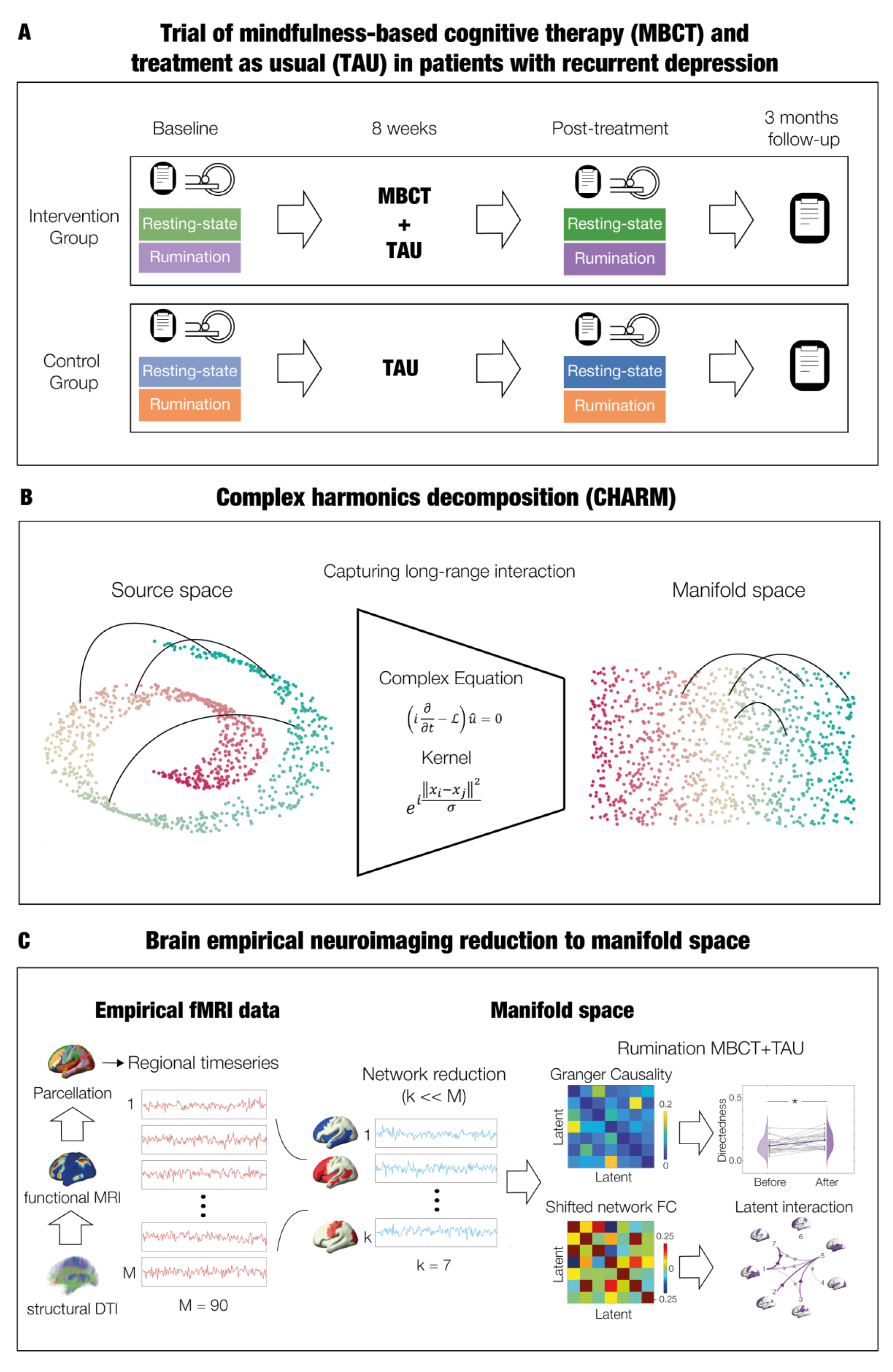
Methodology for extracting low-dimensional networks in brain dynamics before and after treatment for depression with Mindfulness-Based Cognitive Therapy (MBCT). **A.** Design of trial (single-blind randomised controlled study) with two groups of intervention: MBCT plus treatment as usual (TAU) (MBCT+TAU), or TAU alone. Clinical and behavioural assessment, and functional magnetic resonance imaging (fMRI) scans, are performed at baseline and post treatment. **B.** Schematic of the process of decomposition, the neighbourhood of components is indicated with colourmaps and the long-range connections are indicated with lines. The complex harmonics diffusion maps (CHARM) framework uses a complex kernel derived from the Schrödinger Wave equation. This way, local relationships are captured, as well as non-local connections which arise from the underlying criticality and anatomical long-range connections. **C.** Whole-brain dynamics arise from neural activity coupled through the underlying anatomy with short-range wiring and long-range connections. The brain data is measured with fMRI and divided into M brain areas (i.e., parcellated). The regional timeseries is a high-dimensional source space and can be decomposed into low-dimensional networks (e.g., Yeo resting-state networks). Here, we implemented CHARM to obtain the k manifold space. Computations on the reduced space are done such as quantification of the hierarchical organisation of the Gaussian Granger Causality (GC) matrix, and top latent interactions identified in the shifted network Functional Connectivity (FC). Figure adapted from (Deco et al., 2025) under Creative Commons Attribution License 4.0 (CC BY).

### Intervention

The two arms were MBCT+TAU, or TAU alone. Mindfulness-based cognitive therapy is a program of 8-week duration developed to prevent relapse or recurrence to depression (Segal, 2013). The therapy combines elements from cognitive behavioural therapy for depression, and mindfulness meditation from the mindfulness-based stress reduction program. The training was delivered in a university setting by experienced therapists following the MBCT treatment manual (van Aalderen et al., 2015). Therapists had at least 7 years of experience teaching MBCT in Danish based on internationally recognized guidelines for teachers, trainers and supervisors of mindfulness courses (Crane & Kuyken, 2019; Kuyken, 2012). They had the highest rating on the Mindfulness-based Interventions Teaching Assessment Criteria Measure and were supervisors and trainers by other mindfulness teachers (Fresco et al., 2007). The whole program consisted of a pre-class interview, weekly classes (2.25 hours duration each), daily homework, a practice day close to the 6^th^ session, and after the end of the 8-week program 4 booster sessions were offered.

On the other hand, TAU can consist of psychological therapy or antidepressant medication. In this study, it was restricted to either no medication or a stable dose of antidepressant medication, but no psychotherapy. If on medication, participants were encouraged to adhere during the whole trial and if any changes, to report them, without affecting their participation.

Assessment consisted of clinical and behavioural measures and magnetic resonance imaging at baseline before randomisation and within a month after treatment. Depressive symptoms were also assessed at 3 months after randomisation. For the MBCT+TAU group, mean attendance and practice were above 7 (out of 8 sessions) and above 3 days (per week), respectively.

### Clinical measures and psychological processes

The clinical and behavioural measures consisted in: Quick Inventory of Depressive Symptomatology-Self Report (QIDS) for assessing depressive symptoms (Rush et al., 2003); perceived stress with the Perceived Stress Scale (PSS) (Cohen et al., 1983); interoceptive awareness using the subscales of noticing (NO), emotional awareness (EA), body listening (BL), attention regulation (AR), trusting (TR), and not-distracting (ND) of the Multidimensional Assessment of Interoceptive Awareness (MAIA) (Mehling et al., 2012); decentering based on the Experiences Questionnaire – decentering factor (EQ) (Fresco et al., 2007); mindfulness skills using the Five Factor Mindfulness Questionnaire – short version (FFMQ) (Baer et al., 2008; Tran et al., 2013); and trait rumination with the Rumination Response Scale (RRS) (Langenecker et al., 2024; Roelofs et al., 2006).

### fMRI paradigm

The functional magnetic resonance imaging (fMRI) consisted of an initial structural scan and 4 separate consecutive functional scans at different conditions (i.e., states of mind) each lasting 5 minutes. The scan conditions were: resting-state, mindfulness state, resting-state, and rumination state, always in that order. Before each scan, the research team instructed participants the nature of the condition and the voluntary aspect due to ethical motives (participants could opt out). After each scan, an experience sampling was done where participants had to rate their affective, cognitive, and somatic experiences, adapted from (Smallwood et al., 2016). It consisted of a computer screen with a visual analog scale (VAS) from 0-100% and a cursor to move with a trackball. A brief follow-up talk was also done by clinically trained members to make sure participants were fine.

For this analysis, we select as scan conditions the first resting-state and the rumination induction state. The effectiveness of the latter was revealed in the differences from VAS scores showing increased negative self-related thoughts (‘I had negative thoughts about myself’) and decreased body awareness (‘I was aware of my body’) in rumination compared to resting-state in all sessions and intervention arms (shown in (Dagnino et al., 2026)). For more details on MRI paradigm and rationale please see **Supplementary Information**.

### Brain parcellation

Neuroimaging data was parcellated using the Automated Anatomical Labelling (AAL) atlas into 90 brain areas (Tzourio-Mazoyer et al., 2002).

We have the number of voxels of each cortical and subcortical brain area overlapping the 7 well-known Yeo networks (Visual, Somatomotor, Dorsal Attention, Salience, Limbic, Control and Default Mode Networks) (Yeo et al., 2011).

### Complex harmonics decomposition (CHARM)

There are different approaches to reduce brain activity from the regional level (i.e., source space) to a lower dimensional representation (i.e., manifold space). Brain dimensionality reduction allows to describe large-scale brain organisation in terms of a reduced number of meaningful manifolds (i.e., latents), each corresponding to a pattern of coordinated activity across brain regions (i.e., low-dimensional networks). The mathematical formulation of the manifold reduction problem using Laplacian eigenmaps (Belkin, 2003) consists of constructing a similarity graph from empirical data (i.e., graph Laplacian) and performing an eigendecomposition. The resulting eigenvectors define a set of spatial modes that can be used to represent spatial patterns of brain activity. This formulation is mathematically linked to the heat equation, whose Gaussian kernel describes the diffusion on the graph. The approach has been successfully applied in neuroscience using similarity matrices derived from structural connectivity (Atasoy et al., 2016), functional connectivity (Vohryzek et al., 2024) and coherence connectivity dynamics (Rue-Queralt et al., 2021). In this study, we implemented complex harmonics decomposition (CHARM) (Deco et al., 2025), a framework that reformulates the manifold reduction problem using the complex kernel coming from the Schrödinger wave equation, instead of the Gaussian kernel coming from the heat equation (which has similar mathematical structure) (Schleich et al., 2013; Schrödinger, 1926a, 1926b). This formulation allows to capture interferences and aggregation of information transfer mediated by nonlocal brain dynamics arising from criticality and amplified by anatomical long-range connections (**Figure 1B**). Furthermore, in CHARM, the similarity matrix is constructed at the temporal scale. As a result, CHARM provides a set of manifolds evolving over time, each one representing a unique coordination of activity across regions. For full mathematical details of CHARM please see **Supplementary Information**.

### Overview of analytical approach

We applied CHARM to two primary comparisons, rumination MBCT+TAU before vs. after, and resting-state MBCT+TAU before vs. after. Subsequent analyses focused on the rumination condition given its increased sensitivity, while additional comparisons (e.g., in TAU) were implemented to evaluate the robustness of the method. CHARM was applied separately to pairs of groups to facilitate the detection of condition- or treatment-related changes while ensuring balanced sample sizes and limiting computational complexity. In all analysis, we reduced the regional source space (90 brain areas * timepoints) to a manifold space of *k* low-dimensional networks (*k* latents * timepoints) (**Figure 1C**).

After uncovering the low-dimensional networks, we implemented different analyses which are described in detail in the **Supplementary Information**. First, we measured the global hierarchical organisation of each group in the reduced space by quantifying the directedness (Deco, 2024) of the Gaussian Granger Causality (GC) matrices (Granger, 1980). The GC infers the directed predictive interactions between the timeseries of each network in each group. We compared results with classical dimensionality approaches, by calculating the GC on the latents obtained with Yeo resting-state networks (RSN) (Yeo et al., 2011) and principal component analysis (PCA) (Jolliffe & Cadima, 2016). Second, we assessed low-dimensional networks interactions in each group by measuring the time-lagged asymmetry, computed as the shifted network functional connectivity (FC) on the manifold space. We also evaluated the ability of significant interactions to discriminate between groups using a support vector machine (SVM). Third, in most cases, we investigated the nature of each latent by computing the contribution of the source space (i.e., regional timeseries) to the manifold space (i.e., latent timeseries), building functional projection maps, and their overlap with canonical Yeo RSNs. Finally, for the rumination state of mind in the MBCT+TAU group, we identified recurring configurations of latent co-activation using a clustering approach inspired by the leading eigenvector dynamic analysis (LEiDA) method (Cabral et al., 2017).

### Comparisons of brain measures with psychological processes and clinical outcomes

We compared the relation between different brain measures with psychological processes and clinical outcomes using Spearman and partial Spearman correlation corrected by covariates.

### Statistical Analysis

All statistical analyses were done with permutation-based Wilcoxon sign-rank or rank-sum (1000 permutations and 0.05 as significance threshold). At a node-level, we performed Wilcoxon sign-rank or rank-sum and reported the z-value (standardized test statistic), determining significance with the corresponding p-values. We corrected for multiple comparisons, when applicable, implementing False Discovery Rate (FDR) method (Hochberg & Benjamini, 1990).

## Results

We first computed CHARM (Deco et al., 2025) to uncover low-dimensional networks in MDD before and after MBCT+TAU separately for resting-state (N=39 subjects for each session) and rumination (N=26 subjects for each session). Less participants underwent rumination given its voluntary nature for ethical reasons, which had similar baseline characteristics than the participants avoiding rumination except for age and lower depression scores (details on demographics are described in (Dagnino et al., 2026)). We reduced the dimensionality from the source space (90 brain areas * timepoints) to the manifold space (7 latents * timepoints), where each latent (i.e., low-dimensional network) represents a pattern of coordinated brain activity including both short and long-range interactions. We selected a manifold dimension of *k* = 7, as it provided a stable reconstruction of the data (**Supplementary Figure S1**), fewer dimensions would miss relevant patterns, while more would add complexity without meaningful information.

### Global directedness in the manifold space increases for MBCT+TAU during rumination

To begin, we aimed to capture the hierarchical organisation of the manifold space and evaluate whether it aligns with the source space (Dagnino et al., 2026). To do so, we quantified the directedness of causal interactions between the different latent timeseries. We first estimated the directed interactions using Gaussian Granger Causality (GC) (Granger, 1980), which evaluates the contribution of past activity in one manifold to predicting the future activity in another. Then, we quantified the global hierarchy of the reduced space using the measure of directedness (Deco, 2024).

Results showed a significant increase in directedness in the rumination state of mind (p < 0.05) but not in resting-state (**Figure 2A**), consistent with changes in directedness in the regional level (Dagnino et al., 2026). The change in directedness in the manifold space performing dimensionality reduction with the classical Yeo resting-state networks (RSN) (Yeo et al., 2011) or principal component analysis (PCA) did not reproduce the results observed with CHARM.

**Figure 2.**
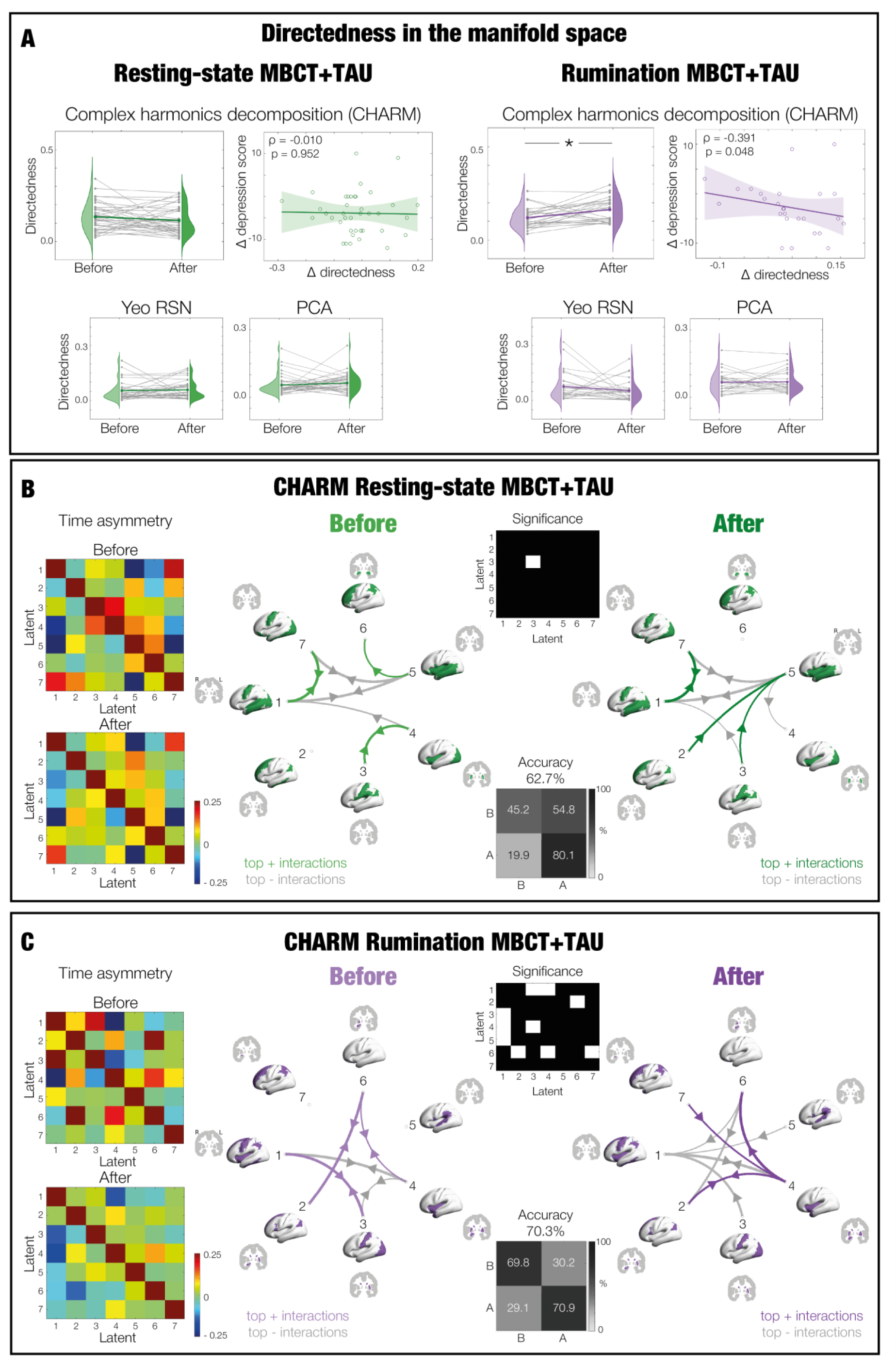
CHARM captures distinct low-dimensional network interactions in MBCT+TAU resting-state and MBCT+TAU rumination. **A.** We calculated the interactions of the latent timeseries using Gaussian Granger Causality (Granger, 1980), the unnormalised form of directed transfer entropy under Gaussian assumptions (Deco, 2021). Then, we computed the directedness (Deco, 2024), showing it changes significantly with treatment in rumination – increasing (* represents p < 0.05) - and not resting-state. This is not captured by the classical Yeo RSN nor principal component analysis (PCA). Grey lines represent the trajectory of each individual whereas the coloured line represents their averaged trajectory. Only in rumination, the positive change in directedness in the CHARM reduced space at a subject level significantly correlates with a negative change in QIDS (reduction in depression symptoms). For both **B.** resting-state MBCT+TAU and **C.** rumination MBCT+TAU, the left matrices show the shifted network functional connectivity between the 7 manifolds before and after treatment, with strength determined with the colourbars. These time asymmetries are schematised (the top 20%) in the circular maps with strongest positive interactions in violet and negative interactions in grey. Arrow thickness represents the strength of the interactions. Significance (p < 0.05) between groups is represented in the white squares of the black and white matrices. Importantly, the schematics depicting top 20% of latent interactions within each group align partially with the significant differences identified by the white squares in the matrices, as these reflect different comparisons (the former shows the within-group asymmetry whereas the latter shows the between group statistical differences). Lastly, we used a support vector machine (SVM) to perform a pattern separation between before and after treatment. In each case (CHARM resting-state and CHARM rumination), we used as input the interactions showing significant differences between the corresponding sessions (before and after).

We then computed the Spearman correlation between the change (after-before) in global directedness in the manifold space with the change (after-before) in clinical and behavioural scores. These were significant for the rumination state of mind and not resting-state, negatively correlated for the depressive symptoms measured with self-reported 16-Item Quick Inventory of Depressive Symptomatology (QIDS) post treatment (rho = -0.391, p = 0.048) (**Figure 2A**). It was also significant, positively correlated, for decentering factor of the Experience Questionnaire (EQ) (rho = 0.524, p = 0.007) and Multidimensional Assessment of Interoceptive Awareness (MAIA) subscales of emotional awareness (EA) (rho = 0.436, p = 0.029), attention regulation (AR) (rho = 0.495, p = 0.012) and body listening (BL) (rho = 0.410, p = 0.042). We repeated the analysis using Partial Spearman correlation corrected for covariates of age, sex and ADM. Significance remained only for QIDS and EQ. Finally, we repeated the analysis using Partial Spearman correlation corrected for the same covariates (i.e., age, sex and ADM), together with baseline QIDS. Significant was lost for all cases. No correction for multiple comparisons was done for either case given the analysis was exploratory (**Supplementary Tables S1 and S2**).

### Low-dimensional network interactions between brain states captured by CHARM

We then obtained the time asymmetric interactions before and after treatment for each condition by calculating the shifted network functional connectivity between the timeseries of each of their corresponding 7 latents. We calculated this in the CHARM reduced space of resting-state (**Figure 2B**) and in the CHARM reduced space of the rumination state of mind (**Figure 2C**). The circular plots with the strongest 20% interactions and the matrices with the significant differences between sessions (before vs. after) show highest changes after treatment compared to baseline in the rumination state of mind and not in resting-state. Specifically, the manifold interaction is more diverse, with an increased proportion of negative interactions. Furthermore, we applied a support vector machine (SVM) to evaluate whether the significant interactions carry sufficient information to discriminate between pre- and post- treatment in each condition. Accuracy levels reached 62.7% for resting-state and 70.3% for rumination.

### CHARM before and after MBCT+TAU during rumination

Given the increased sensitivity of the rumination state of mind in capturing MBCT+TAU changes, we focused on this scan condition for the rest of the analysis.

#### Characterisation of each latent

To understand how each latent is related with regional activity, we computed the functional projection of brain dynamics from the source space to each latent in the manifold space specifically for the CHARM analysis of rumination before and after MBCT+TAU. By calculating the correlation between the timeseries of each brain area in the source space, with the timeseries of each latent in the manifold space, we could identify how strongly each brain region contributes to a given latent over time. In other words, from a source matrix [NxT] and projected matrix [MxT] (where N are brain regions, M are latent dimensions and T are timepoints), we built a functional projection map [NxM].

For each latent, we rendered the contribution of cortical and subcortical brain areas in **Figure 3A**, and the strongest positive and negative values are listed in **Supplementary Table S3**. Overall, latents where characterised by widespread contributions across areas, with the majority containing areas in their top or bottom extremes related with bodily (e.g., precentral and postcentral gyrus) and interoceptive processing (e.g., insula, anterior cingulate cortex). In particular, latents 1 and 4 had an overall antiphase relationship between source brain dynamics and manifold networks (i.e., temporal fluctuations of all brain areas presented an inverse projection to the latent). In contrast, latent 2 had all positive phases, led by subcortical (e.g., caudate, putamen, hippocampus) and frontomedial regions (e.g., anterior cingulate cortex, medial superior frontal cortex). The rest of the latents had some areas in phase and others in antiphase. Latent 3 exhibited positive values for visual (e.g., occipital cortex, cuneus) and somatomotor areas (e.g., postcentral gyrus, paracentral lobule). Latent 5 was led by limbic (e.g., rectus gyrus, olfactory cortex) and frontomedial regions (e.g., orbitofrontal cortex, anterior cingulate cortex) whereas latents 6 and 7 by temporal (e.g., temporal cortex, heschl gyrus), sensorimotor (e.g., precentral gyrus, supplementary motor area) and frontoparietal areas (e.g., inferior parietal cortex, middle frontal cortex).

**Figure 3.**
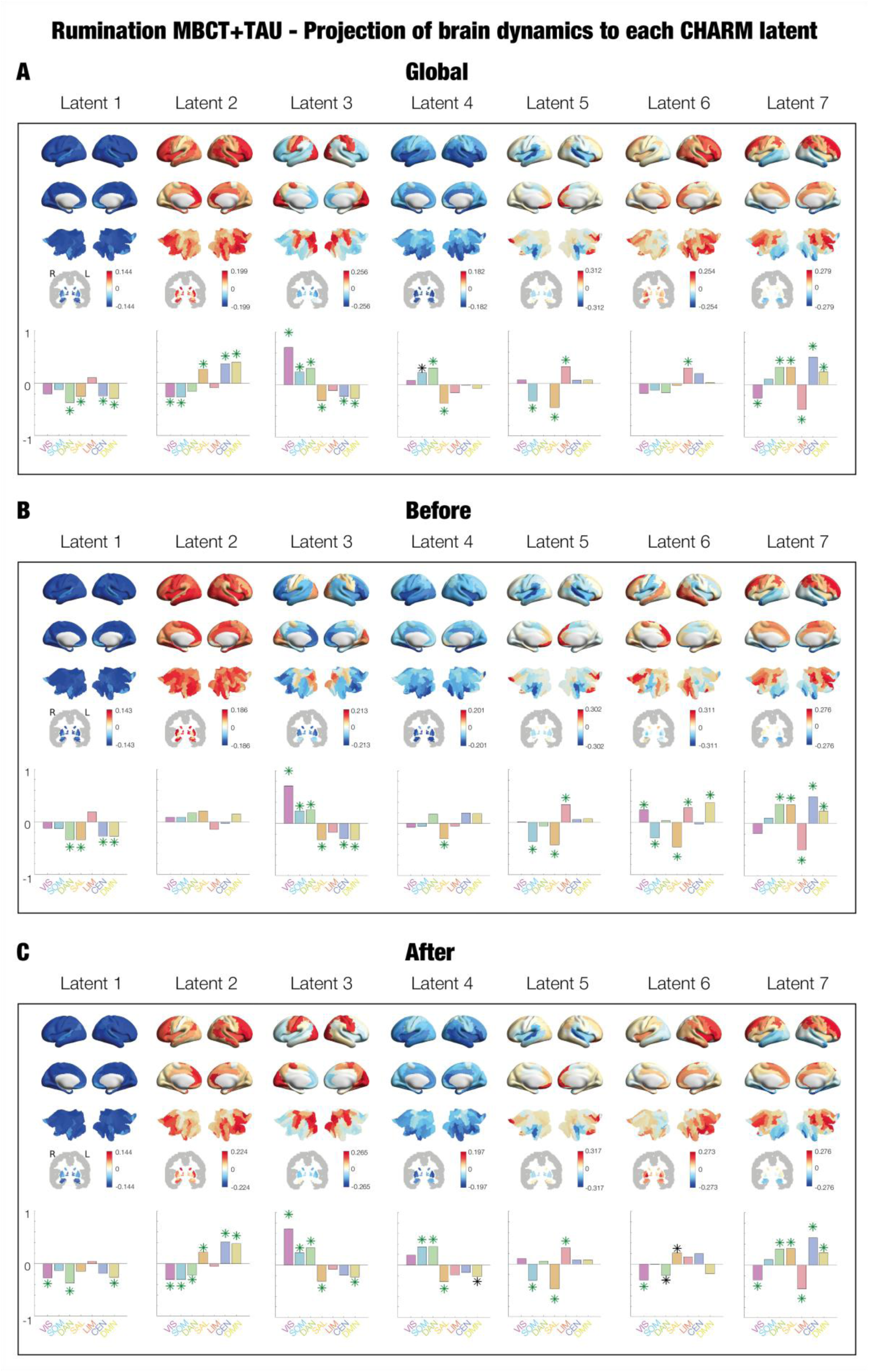
Rumination MBCT+TAU projection of brain dynamics into each manifold network. For the CHARM manifold space, the brain renders show the contribution of the whole-brain dynamics in the source space to each latent of the reduced space (i.e., projection map). We also calculated the correlation between these contributions and the prototypical Yeo RSN. We built this for the **A.** global reduced space, **B.** before treatment and **C.** after treatment. Subcortical areas are shown on slices in Montreal Neurological Institute (MNI) space (coronal axis y=-6 mm).

Furthermore, to evaluate whether the low-dimensional networks were confined to predefined canonical Yeo RSNs (Yeo et al., 2011), we computed the overall spatial pattern alignment of the projection map of each latent with each RSN. To do so, we built a Yeo RSN layout with the proportion of voxels of each brain area to each network (matrix of size brain areas * networks). Then, for each latent, and for each network, we computed the correlation between the functional projection map of the latent, with the proportion of voxels assigned to the network from the RSN layout. Results showed that each latent overlaps with several Yeo RSN, indicating that the manifolds capture distributed network organisation specific to rumination across sessions in MBCT for depression (**Figure 3A**).

Given that the projection map is built using both sessions (before and after), we built new projection maps for each session separately and calculated their significant differences (**Figure 3B-C, Supplementary Figure S2 and Table S4**). Latent 3 showed almost all 90 AAL brain areas with significant differences after treatment, surviving correction by multiple comparisons. Latent 4 presented 50% of areas significantly different, with 20 areas surviving correction by multiple comparisons. Latents 1 and 5 contained approximately 10-15% areas with significance, with 2 and none areas surviving correction by multiple comparisons. Lastly, the rest of the latents (latents 2, 6 and 7) contained very few (e.g., 2 areas) or no significant differences, non-surviving correction for multiple comparisons.

#### Latent configuration in manifold space and their clinical and behavioural relevance

The low-dimensional networks are uncovered in the time domain, therefore to better understand how these patterns evolve, we asked whether different latent activities combined in recurring configurations. The purpose was to capture the dynamic reorganisation of the low-dimensional networks and assess how they differ across sessions and relate to clinical and behavioural outcomes. To address this, we applied a clustering approach inspired by leading eigenvector dynamic analysis (LEiDA) (Cabral et al., 2017). Specifically, we clustered all observables (i.e., timepoints) from the manifold space, each containing the activity of the 7 latent dimensions of the manifold space. This allowed us to identify a set of recurring configurations of latent co-activations (i.e., manifold metastates). For each subject, we then quantified the probability of being in each of these metastates. We applied the K-means algorithm for the number of centres *c*=2-10. The centres yielding maximum proportion of significant differences between sessions, surviving correction for multiple comparisons, were *c*=3 and *c*=7 (**Supplementary Figure S3**). We chose *c*=7 given it revealed a more balanced fractional occupancy between clusters.

The latent coactivation for each centroid is represented in **Figure 4A**. We then calculated the probability of occurrence for each subject in each session (before and after) (**Figure 4B**). Results showed a significantly lower probability of occurrence after treatment of metastates C and F (p < 0.05), both governed by latent 5 negatively and positively, respectively. Furthermore, there was a significantly higher probability of occurrence after treatment of metastates D and E (p < 0.05), both led by latents 3 and 6. In particular, metastate D was led positively by both latents, whereas metastate E was led negatively by latent 3 and positively by latent 6. We also obtained the transition probability matrix for each session and calculated the entropy, where higher entropy indicates more frequent and diverse state transitions. Results showed significant increased entropy following treatment (**Figure 4C**).

**Figure 4.**
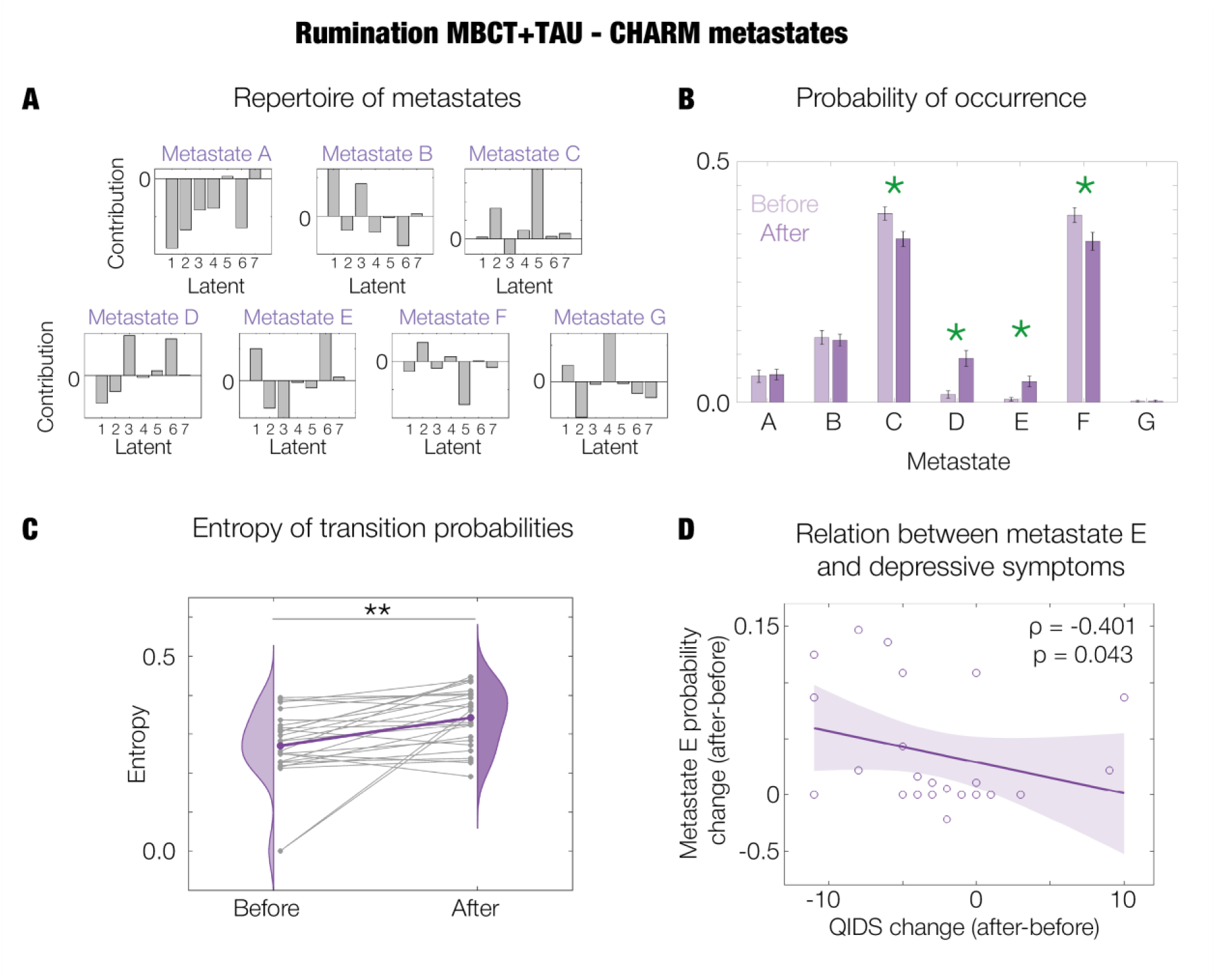
Clustering latent observables of rumination MBCT+TAU into metastates. **A.** Each cluster centre represents a metastate, which is a combination of the 7 latent networks. **B.** Probability of occurrence of each session (before and after) in each metastate. **C.** Change in entropy of transition probability matrices during rumination following MBCT+TAU. Grey lines represent the trajectory of each individual whereas the coloured line represents their averaged trajectory. **D.** Spearman correlation between change in probability of occurrence of metastate E and change in depression scores. Statistics are represented by asterisks in the corresponding plots (*, p < 0.05; **, p < 0.01).

Finally, we calculated the relation between the change (after-before) in probability of occurrence of each metastate with the change (after-before) in clinical and behavioural scores. The Spearman correlation, Partial Spearman correlation correcting for covariates (age, sex and ADM), and the Partial Spearman correlation correcting for the aforementioned covariates together with baseline QIDS can be found in **Supplementary Figure S4**. No correction for multiple comparisons was done given this analysis was exploratory. For the Spearman correlation, most significance was found for Metastates D and E, with increases in probability of occurrence related with improvement in clinical and behavioural outcomes, and Metastate F, with the opposite trend. Specifically, Metastate D was significant for mindfulness skills using the FFMQ (rho = 0.509, p = 0.009), perceived stress measured with PSS (rho = -0.537, p = 0.006), decentering factor of the EQ (rho = 0.561, p = 0.003) and MAIA subscale of AR (rho = 0.521, p = 0.008). For Metastate E it was significant for depressive symptoms measured with QIDS post treatment (rho = -0.401, p = 0.043) (**Figure 4D**), mindfulness skills using the FFMQ (rho = 0.477, p = 0.016), perceived stress measured with PSS (rho = -0.486, p = 0.014), decentering factor of the EQ (rho = 0.489, p = 0.013), and MAIA subscale of AR (rho = 0.491, p = 0.013). For Metastate F it was significant for depressive symptoms measured with QIDS at 3 months follow-up (3M-FU) (rho = 0.423, p = 0.031), mindfulness skills using the FFMQ (rho = -0.507, p = 0.010), decentering factor of the EQ (rho = -0.676, p < 0.001), and MAIA subscales of AR (rho = -0.646, p < 0.001) and BL (rho = -0.470, p = 0.018).

#### Complementary measures

As an exploratory analysis, we computed the irreversibility in the time-asymmetries of the reduced space revealing overall increases after treatment (**Supplementary Figure S5**). Moreover, we measured the contraction of the reduced space, by first calculating the centre of the reduced space (i.e., barycentre) considering all of the 7 latents, and measuring the distance of each timepoint to the barycentre. Then, for each subject, we averaged the distances of the timepoints of the corresponding scanning session. Results showed significant increases with treatment (**Supplementary Figure S6**). Lastly, other measures calculated for each latent in the manifold space (hierarchical position, irreversibility, and global network brain connectivity) presented at least one significant difference in latents 1, 2, 3 and 6 (**Supplementary Figure S7**). All methods are described in detail in the **Materials and Methods in Supplementary Information**.

### CHARM works well with other brain states

We then evaluated the efficacy of CHARM in revealing low-dimensional networks specific to other brain states, and evaluated differences between scan conditions as well as changes for the TAU group. First, we performed CHARM combining the two conditions (resting-state and rumination) at baseline (for the two intervention arms together, MBCT+TAU and TAU) and studied depression manifestations pre-treatment (N=47 subjects for each condition, corresponding to the subjects that performed both resting-state and rumination). Second, we performed CHARM to the conditions post-treatment for the MBCT+TAU group to study differences between resting-state and rumination after mindfulness training (N=26 subjects for each condition, corresponding to the subjects that performed both resting-state and rumination). Third, we computed CHARM in the TAU group of the clinical trial for resting-state (N=26 subjects for each session) and rumination (N=21 subjects for each session), separately. As a note, participant baseline psychiatric variables and questionnaire scores and demographics were similar between the two intervention arms (details provided in (Dagnino et al., 2026)). The reconstruction errors reached stability in all cases using the same parameters and dimensions from the main analysis (**Supplementary Figure S8**).

At a global level, the directedness in the manifold space revealed significant differences between conditions for post-treatment MBCT+TAU (p < 0.01), higher for rumination compared to resting-state, and between sessions for rumination TAU (p < 0.05), increasing after treatment (**Supplementary Figure S9**). Significance was lost when calculating the directedness when reducing the dimensionality with Yeo RSN or PCA. The rest of the comparisons (baseline resting-state vs. rumination, resting-state TAU before vs. after, and rumination TAU before vs. after) did not present significant changes for either analysis (i.e., CHARM, Yeo RSN, PCA).

Focusing on the first and second CHARM analysis of this section, the differences between conditions (resting-state and rumination), there were more significant differences in low-dimensional network interactions post-MBCT+TAU compared to baseline (**Supplementary Figure S10**). Furthermore, discriminative accuracies of the SVM for pattern separation increased from 68.9% before treatment to 83.2% after treatment. With respect to the projection maps, these were unique to each CHARM analysis, with some overlaps between baseline and post-treatment (**Supplementary Figures 11A and 12A**). The differences between conditions were relatively sparse and fragmented at baseline (**Supplementary Figure 11B**), and more spatially continuous, larger and significant post-treatment (**Supplementary Figure 12B**).

Regarding the third CHARM analysis of this section, the TAU arm, the top 20% of interactions were very similar between before and after treatment within each condition, with almost no significant differences (**Supplementary Figure S13**).

## Discussion

In this novel secondary analysis of mindfulness based cognitive therapy (MBCT) for major depressive disorder (MDD), we successfully implemented complex harmonics decomposition (CHARM) (Deco et al., 2025) to understand how mindfulness targets depressive ruminative processing (**Figure 1**). Our results showed distinct distributed spatiotemporal low-dimensional networks across brain states and outperformed traditional dimensionality reduction techniques. During rumination after MBCT treatment manifolds consistently recruited regions involved in bodily and interoceptive processing integrated within the whole-brain and showed significant differences between sessions. We also found latent reconfigurations associated with clinical and behavioural improvements, and greater brain flexibility within the reduced space. To our knowledge, this is the first study implementing CHARM to investigate treatment-related brain changes in a neuropsychiatric disorder, specifically MBCT for MDD. We built on and extended our previous data-driven network approach (van der Velden, 2025) and brain hierarchy study (Dagnino et al., 2026) by uncovering low-dimensional manifold networks in time underlying brain activity, and in turn enabling the quantification of hierarchical organisation within the reduced space.

Our work highlights the relevance of distributed network-level brain computation beyond locality, supported by low-dimensional manifolds capturing local and non-local interactions in critical brain dynamics captured by CHARM (Deco et al., 2025), to study the neural processes through which MBCT exerts its therapeutic effects in MDD. We uncovered distinct low-dimensional manifold networks across analyses, demonstrating flexibility and brain state specificity. Importantly, the low-dimensional organisation was free from assumptions on pre-defined networks (e.g., one-to-one mapping to static networks), enabling us to characterise brain activity specific to a given brain state (**Figure 2B-C, Supplementary Figures S10 and S13**). Furthermore, manifolds were obtained in the spacetime domain, allowing us to study dynamical interactions occurring within the low-dimensional space. Finally, CHARM showed greater efficacy compared to traditional dimensionality techniques (Yeo RSN and PCA) in terms of the ability to capture hierarchical brain organisation (**Figure 2A and Supplementary Figure S9**).

Interestingly, our results on brain hierarchy in the reduced space are closely aligned with our past investigation in the regional space (Dagnino et al., 2026), showing that the hierarchical brain organisation unfolds on a low-dimensional manifold embedded within the high-dimensional space. Specifically, we found significant increases of brain hierarchy during rumination following MBCT treatment, significantly correlated with a reduction in depressive symptoms and increased decentering, emotional awareness, attention regulation and body listening. No change was observed during resting-state following MBCT+TAU (**Figure 2A and Supplementary Table S1**). Furthermore, during rumination MBCT+TAU, the manifold with most significant differences (latent 3) (**Supplementary Figure S2**) followed the well-known cortical gradient-like organisation with low-order areas in one extreme and higher-order areas in another extreme (Bernhardt et al., 2022; Huntenburg et al., 2018; Margulies et al., 2016). This regional organisation showed stronger expression post-treatment compared to baseline, indicating more sustained temporal expression and stability. In our previous investigation in the regional space (Dagnino et al., 2026), a higher global brain hierarchy together with an increased regional differentiation following treatment during rumination was related with clinical and behavioural improvements. These neural changes were interpreted as a possible underlying brain process of disengagement from locked ruminative cycles following mindfulness training. Given that increased hierarchical brain organisation following mindfulness-training is observed across different scales (i.e., from brain regions to low-dimensional manifolds), we suggest that the effect is robust and reflects a core property of whole-brain re-organisation following treatment.

We focused on rumination given its increased sensitivity compared to resting-state in unravelling neural changes following MBCT+TAU (**Figure 2**), showing that a brain state directly related to the disorder provides substantially richer neural information, in line with the previous studies in the same cohort (Dagnino et al., 2026; van der Velden, 2023). Our results on the projection of brain dynamics from the source space to each latent of the manifold space and differences between sessions during rumination uncovered low-dimensional manifolds distributed across multiple interacting systems, with non-local interferences and aggregation of information. The manifolds recruited distributed cortico-limbic and self-referential systems involved in cognitive, emotional, and internal processes (**Figure 3**). Furthermore, regions associated with interoceptive and bodily processing were consistently represented across multiple latent dimensions, consistent with our previous analyses in the same dataset (Dagnino et al., 2026; van der Velden, 2023; van der Velden, 2025). Here, we showed their integration into the whole-brain distributed configurations underlying depression response to mindfulness treatment.

We speculate that the integration of interoceptive and bodily processing systems within whole-brain network organisation may facilitate the incorporation of ongoing sensory and interoceptive experience into distributed brain processing following mindfulness training. The interoceptive processing regions, related to the awareness of internal bodily states, go in line with past findings of psychological treatments for depression (Downar et al., 2016; Godlewska et al., 2018; Young et al., 2018). Previous research has found that interoceptive awareness facilitates decentering from rumination and mediates reduced depressive symptoms following a mindfulness-based intervention (Fissler, 2016), whereas experiential avoidance is related to depression vulnerability (Barnhofer et al., 2014). On the other hand, recent work showed that self-generated thought can be strongly embodied, which, although associated with negative affect in the moment, has been linked to lower depressive symptoms (Banellis et al., 2026). Considering this, our findings may indicate that decentering is not limited to thoughts but encompasses the whole mind-body state, enabling the participant in MBCT to develop the ability to experience thoughts and feelings as transient mental events and to step back from ruminative thinking patterns by being more aware of and anchored in present-moment embodied experience.

Given that CHARM uncovers manifolds in time, we built a repertoire of coactivation of latents (i.e., metastates) in the reduced space we then associated the changes in probability of occurrence with changes in clinical and behavioural outcomes, reflecting treatment-related latent reconfigurations related with improvements (**Figure 4 and Supplementary Figure S4**). Our results showed increased entropy following treatment (**Figure 4C**), quantifying greater diversity in the transition probabilities between latent configurations and reflecting increased brain flexibility. This was also supported by greater dispersion of timepoints in the reduced space (**Supplementary Figure S6**). Our results may reflect higher metastable dynamics, defined as the tendency to continuously switch between different states enabling a rich exploration of the brain’s functional repertoire, thought to be facilitated in criticality (Deco & Kringelbach, 2016) (Deco & Jirsa, 2012)Interestingly, depression has been associated with increased temporal stability and less metastability (Vohryzek et al., 2022), increased synchronisation and temporal stability compared to healthy controls (Demirtas et al., 2016), with rigid regimes related to rumination (Zimmern, 2020). Therefore, we speculate that increased brain flexibility may reflect a shift away from rigid repetitive trapped brain configurations towards more flexible functional brain configurations following mindfulness training.

Our work has some limitations worth mentioning. As previously described, the study design means the findings are limited in specificity by the choice of a non-specific control group (TAU), in generalisability to the participants who completed both rest and rumination sessions, and the durations of the fMRI scanning sessions (Dagnino et al., 2026, 2026; van der Velden, 2023; van der Velden, 2025). With respect to the CHARM framework (Deco et al., 2025), it has been implemented until now only in fMRI data, which has the limitation of poor temporal resolution and hence might miss relevant electrophysiological dynamics. Future work could extend the framework to other brain testing modalities.

Overall, building and extending on previous work, we found low-dimensional brain organisation changes in rumination after MBCT treatment for depression. Our results add new layers of understanding including the integration within the whole-brain of regions related with body awareness and interoception, and brain flexibility, which may play a role in the reduced “stickiness” to ruminative thinking patterns. Together, these findings position CHARM as a promising tool for uncovering treatment-related changes in low-dimensional organisation underlying brain activity in neuropsychiatric disorders such as MDD.

## Data availability

Data can be requested upon reasonable request to the corresponding authors.

## Code availability

All analysis was performed in MATLAB R2022a and R2024a (MathWorks; Natick, MA, USA). FSL is available at https://fsl.fmrib.ox.ac.uk/fsl/fslwiki/FslInstallation. Subcortical images were built in Python V3.12. The code is available from the corresponding authors upon request.

## Acknowledgements

This research was funded by a Carlsberg Foundation Internalization Fellowship (CF21_0645) to A.M. P.D. is supported by the Department of Research and Universities of the Government of Catalonia, Agency for Management of University and Research Grants (AGAUR), FI-SDUR programme (Grant No. 2022 FISDU 00229). H.G.R. is supported by grants from the Hersenstchting, ZonMW, Parkinson Foundation, and Radboudumc. He obtained unrestricted educational grants from J&J and served as a speaker for J&J, Lundbeck, E-wise, Prelum and Benecke. Y.S.P. and J.V. and G.D. are supported by the EU funded Project NEurological MEchanismS of Injury, and Sleep-like cellular dynamics (NEMESIS; ref. 101071900) funded by the EU ERC Synergy Horizon Europe. Y.S.P. is also supported the Grant PID2024-162576NA-I00 funded by MICIU/AEI/10.13039/501100011033 and by “ERDF A way of making Europe”, ERDF, EU. G.D. is also supported by Grant PID2022-136216NB-I00 funded by MICIU/AEI/10.13039/501100011033 and by “ERDF A way of making Europe”, “ERDF, EU”, AGAUR research support grant (ref. 2021 SGR 00917) funded by the Department of Research and Universities of the Generalitat of Catalunya, and Grant PID2024-155136NI-I00 financed by MICIU/AEI/10.13039/501100011033/ and by “ERDF A way of making Europe”, ERDF, EU. M.L.K. is supported by the Centre for Eudaimonia and Human Flourishing (funded by the Pettit and Carlsberg Foundations) and Center for Music in the Brain (funded by the Danish National Research Foundation, DNRF117).

