## Supplementary Information for "Complex harmonic manifolds in mindfulness-based cognitive therapy for major depressive disorder"

### Materials and Methods

#### - Participants

The participants were recruited in general practices and psychiatric units in Central Jutland region, Denmark. Inclusion criteria consisted in: diagnosis of recurrent major depressive disorder, with or without a current episode, with the Structured Clinical Interview for DSM-IV-TR (Gorgens, 2011); a number of 3 or more previous major depressive episodes; if on antidepressant medication, a stable dose during a minimum of 8 weeks of selective serotonin re-uptake inhibitor or serotonin and norepinephrine reuptake inhibitor; 18 years old or older. Exclusion criteria consisted in: current severe major depressive episode (Beck Depression Inventory-II > 28 (Beck, 1996)); previous completion of MBCT/mindfulness-based stress reduction program; concurrent formal psychotherapy; exhaustive meditation experience (i.e., regular practice, retreats); current severe substance abuse, organic mental disorder; current/past psychosis, pervasive development delay; persistent antisocial behaviour, self-injury; history of schizophrenia, schizoaffective disorder, bipolar disorder; antipsychotic medication; benzodiazepines. All participants gave written informed consent.

#### - Randomisation and blinding

Randomisation of participants was done in a computerised system by an independent researcher, which allocated them in a 5:3 ratio to an 8-week MBCT+TAU training, or TAU. Participants were stratified based on symptoms using Beck Depression Inventory-II (Beck, 1996) and antidepressant usage to balance across the two arms the distribution of patients with and without symptoms and antidepressants. Treatment allocation was masked at baseline but not afterwards for participants, therapist and trial coordination. Clinical interviews assessing relapse risk and MRI scans were masked during the whole trial for researchers.

#### - Instructions for experimental conditions

During the different resting-state scans, participants were told to first relax and close their eyes. This first condition (i.e., state of mind) was chosen in order to assess the general vulnerability of depression without any potential confounding carry-over effects from other conditions. Then, a second mindfulness condition and a third resting-state scan were made, not reported here. During the fourth condition, the rumination state, participants were guided through a rumination induction with closed eyes, and this task was chosen as it addresses depressive rumination. Specifically, participants had to first rehearse a sad autobiographical memory. Then, they had to stay in the sad mood, reflecting on self-related causes and consequences of it. The choice of memory was not restricted over sessions. This paradigm is known to induce negative self-related thoughts in individuals with a history of recurrent depression and has already been validated (Karl et al., 2018).

#### - Magnetic resonance imaging acquisition

A structural three-dimensional T1-weighted (3D-T1) scan was acquired with the following parameters: 176 slices covering the whole brain; echo time (TE)= 3.8 ms; repetition time (TR)= 2300 ms; inversion time=31260 ms; flip angle=8°; field of view (FOV)= 256 mm; spatial resolution=1x1x1 $mm^3$; Generalized Autocalibrating Partially Parallel Acquisitions (GRAPPA)= 2, phase-encoding direction= AP. Functional data was collected in all conditions (resting-state, mindfulness, resting-state and rumination induction) obtaining for each 203 volumes of 2D gradient-echo EPI fMRI data with the following parameters: 52 ascending axial slices covering the whole brain; 3.8 x 3.8 x 3.8 mm³; FOV=192; GRAPPA= 2; Multiband= 2; TE=30 ms; TR=1480 ms; flip angle=65°; phase-encoding direction=AP.

**- BOLD fMRI pre-processing**

The pre-processing was performed with FSL tools (version 6.0) (Smith et al., 2004) using standard procedures. The following were included: skull-stripping (BET tool (Smith, 2002)); functional to structural image registering (FLIRT tool (Jenkinson et al., 2002) with default settings for Boundary-Based registration); structural image to standard space registering (FNIRT tool with default settings, 12 degrees of freedom and 10 mm warp-resolution); motion correction (MCFLIRT tool (Jenkinson et al., 2002)); 5mm kernel for spatial smoothing. Motion correction was performed using an independent component analysis-based strategy for Automatic Removal of Motion Artifacts (ICA-AROMA (Pruim et al., 2015)). The first five eigenvariates of time courses were extracted from white matter and cerebrospinal fluid masks (segmentation done with FAST tool (Zhang et al., 2001)) for further denoising (using *fsl_glm*). Lastly, data was high-pass filtered (100 seconds cut-off).

**Complex harmonics decomposition (CHARM)**

Here we describe the reduction of high-dimensional source space composed by neuroimaging timeseries into low-dimensional manifold networks using complex harmonics decomposition (CHARM). First, $x_i \in \mathcal{R}^M$ is defined, the column vector which contains BOLD signals of the *M* brain regions at timepoint i[th] of the timeseries. Thus, $X = [x_1, x_2, \dots, x_N] \in \mathcal{R}^{M \times N}$ is the matrix of all brain regions spanning a time window of *N* timepoints (i.e., the columns). Assuming brain dynamics lie on a sufficiently smooth low-dimensional manifold (dimension $k$ << *M*) embedded in the high-dimensional $\mathcal{R}^M$ space, the generic problem of dimensionality reduction is defined as follows. Given a set $X$, find a set of points $Y = [y_1, y_2, \dots, y_N] \in \mathcal{R}^{k \times N}$ such that $y_i \in \mathcal{R}^k$ "represents" $x_i$.

The formulation of manifold reduction in continuum space allows for the analytical derivation in discrete space (Belkin, 2003). In continuum space: Let $\mathcal{M}$ be a smooth, compact, *m*-dimensional Riemannian source space (Rosenberg, 1997). The one-dimensional reduction corresponds to a manifold that is a real line, defined in a way that points close together in the source space are mapped close together on the manifold line. Let $f$ be a map from the source space to a manifold line, $f : \mathcal{M} \rightarrow \mathcal{R}^1$ which is twice differentiable. This map can be found by minimizing the following cost function $\mathfrak{H}$, as shown by Belkin and Niyogi:

$$\mathfrak{H} = \int_{\mathcal{M}} \|\nabla f(x)\|^2 dx. \quad (1)$$

The equivalent to minimizing the objective function $\mathfrak{H}$ is finding the eigenfunctions of the Laplace Beltrami operator $\mathcal{L}$, defined by $\mathcal{L}f = -div\nabla(f)$. The eigenfunctions of $\mathcal{L}$ are denoted by $f_i$, where the first eigenfunction is a trivial constant which maps the entire manifold to a single point, and the first nontrivial eigenfunction defines the map to a line. The general case corresponds to the map defining the dimensionality reduction conserving neighbourhood as follows:

$$x \rightarrow [f_1(x), f_2(x), \dots, f_k(x)]. \quad (2)$$

Belkin and Niyogi showed the Laplace Beltrami operator on differentiable functions on a source space $\mathcal{M}$ is intimately related to heat flow (Belkin, 2003). They derived the harmonic decomposition from the heat equation with the partial differential equation $\left(\frac{\partial}{\partial t}+\mathcal{L}\right)u=0$ and solution $u(x,t)=\int_{\mathcal{M}} H_t(x,y)f(y)\,dy$. Here, $H_t(x,y)$ is the heat kernel, the Green's function for the partial differential equation. The initial heat distribution is $u(x,0)=f(x)$ and thus

$$\mathcal{L}f(x)=\mathcal{L}u(x,0)=-\lim_{t\to 0}\frac{\partial}{\partial t}\left[\int_{\mathcal{M}} H_t(x,y)f(y)dy\right]. \tag{3}$$

As the kernel of the heat equation is approximately Gaussian,

$$H_t(x,y)\approx(4\pi t)^{-k/2}e^{-\frac{\|x-y\|^2}{4t}}. \tag{4}$$

As *t* tends to 0, the heat kernel $H_t(x,y)$ tends to Dirac's function $\delta$, $\lim_{t\to 0}\left[\int_{\mathcal{M}} H_t(x,y)f(y)dy\right]=f(x)$. As such, from the definition of the derivative, for small t,

$$\mathcal{L}f(x)\approx\frac{1}{t}\left[f(x)-(4\pi t)^{-M/2}\int_{\mathcal{M}} e^{-\frac{\|x-y\|^2}{4t}}f(y)dy\right]. \tag{5}$$

This expression can be approximated with the following if $[x_1,x_2,\dots,x_N]$ are datapoints on $\mathcal{M}$:

$$\mathcal{L}f(x_i)\approx\frac{1}{t}\left[f(x_i)-\frac{1}{N}(4\pi t)^{-\frac{M}{2}}\sum_{x_j} e^{-\frac{\left\|x_i-x_j\right\|^2}{4t}}f(x_j)\right]. \tag{6}$$

And, since the global coefficients do not affect the eigenvectors of the discrete Laplacian, it can be rescaled such that the eigenfunctions of $\mathcal{L}$ are discretized as the graph Laplacian, as shown by Belkin and Niyogi:

$$\mathcal{L}f(x_i)\approx\sum_{x_j} e^{-\frac{\left\|x_i-x_j\right\|^2}{4t}}f(x_j)-\left(\sum_{x_j} e^{-\frac{\left\|x_i-x_j\right\|^2}{4t}}\right)f(x_i). \tag{7}$$

The nonlinear projection of a point $x_i$ in the reduced manifold latent *k*-dimensional space spanned by the first *k* eigenvectors $[\varphi_1,\ \varphi_2,\dots,\varphi_k]$ of the graph Laplacian has coordinates given by:

$$y_i=[\lambda_1\varphi_1(i),\ \lambda_2\varphi_2(i),\dots,\lambda_k\varphi_k(i)]\quad i=1\dots N. \tag{8}$$

Here, $\varphi_j(i)$ is the $ith$ element of the eigenvector $\varphi_j$. The spectral gap in the eigenvalues of the final decomposition can determine the embedding dimension, providing the classic harmonic decomposition. Overall, the origin of the Gaussian kernel can be analytically derived from the definition of the manifold reduction problem in the continuum space by solving the heat equation when time tends to zero. A direct interpretation of the Gaussian kernel functioning is the implementation of the manifold reduction conserving the discrete neighbourhood space, which is the basis of the harmonic decomposition in **Equation 8**.

Inspired by Schrödinger, in CHARM a complex kernel is developed for capturing interferences and aggregation of information transfer mediated by long-range functional interactions in the brain. Instead of using the heat equation, which is only defined in real space, the Schrödinger equation is used (Schrödinger); the partial differential equation $\left(i\frac{\partial}{\partial t}-\mathcal{L}\right)\hat{u}=0$, which corresponds to the case of a free particle. This way, it can capture non-local effects using a complex equation with time going to zero from the imaginary axis, rather than just from the real axis as with the heat equation. Schrödinger's equation is simplified by using the constraints $\hbar/2m$=1, and the solution is given by $\hat{u}(x,t) = \int_{\mathcal{M}} \widehat{H}_t(x,y) f(y) dy$. The free particle Schrödinger kernel is given by $\widehat{H}_t(x,y)$, known as Green's function for this partial differential equation.

The same steps as above are applied starting from Schrödinger's equation and the results are reproduced using wick transformation $t \rightarrow it$. The discretisation of the eigenfunctions of $\mathcal{L}$ from Schrödinger perspective is

$$\mathcal{L}f(x_i) \approx \sum_{x_j} e^{i\frac{\|x_i - x_j\|^2}{4t}} f(x_j) - \left(\sum_{x_j} e^{i\frac{\|x_i - x_j\|^2}{4t}}\right) f(x_i). \tag{9}$$

The approximation of the graph Laplacian matrix is with a new complex kernel for the matrix $\widehat{W} \in \mathcal{R}^{N \times N}$, whose elements are given by

$$\widehat{W}_{ij} = e^{i\frac{\|x_i - x_j\|^2}{\sigma}}. \tag{10}$$

Here, $\|\ \|$ computes the distance between two points using the Euclidean L2 norm, and $\sigma$ is a scale parameter of the kernel ($\sigma = 300$).

Given that it is more convenient to work with the transition probability matrix, a two-step procedure is used: define the *t*-steps diffusion matrix by taking the power *t* of the diffusion matrix $\widehat{W}^t$ ($t = 3$) and define the nonnormalised probability transition matrix by the square module of the *t*-steps diffusion matrix similar to Schrödinger (Rosenberg, 1997):

$$\hat{Q}(t) = \left|\widehat{W}^t\right|^2, \tag{11}$$

where $|\ \ |$ notates the module.

The diagonal normalization matrix $\widehat{D}$ is defined as follows:

$$\widehat{D}_{ii} = \sum_{j=1}^{N} \hat{Q}_{ij}. \tag{12}$$

And, the normalised transition probability matrix $\hat{P}$ by:

$$\hat{P}(t) = \widehat{D}^{-1}\hat{Q}(t). \tag{13}$$

Singular value decomposition is applied on $\hat{P}$ to get:

$$\hat{P}(t) = \hat{\psi}\hat{\Lambda}\widehat{\psi^T}. \tag{14}$$

Here, $\hat{\Lambda}$ is the diagonal matrix storing the *M* eigenvalues ($\hat{\lambda}_0 = 1 \geq \hat{\lambda}_1 \geq \hat{\lambda}_2$ ..., with the first being the trivial eigenvalue that is equal to 1 given that P is a Markovian matrix). Furthermore, $\hat{\psi} = [\hat{\varphi}_1, \hat{\varphi}_2, \dots, \hat{\varphi}_N] \in \mathcal{R}^{N \times N}$, whose columns $\hat{\varphi}_i$ are the eigenvectors of $\hat{P}$.

The CHARM manifold reduction $\hat{y}_i$ is given by the coordinates of the nonlinear projection of a point $x_i$ in the reduced manifold latent *k*-dimensional space spanned by the first *k* eigenvectors $[\hat{\varphi}_1, \hat{\varphi}_2, \dots, \hat{\varphi}_k]$ of the matrix $\hat{P}(t)$:

$$\hat{y}_i = \left[\hat{\lambda}_1\hat{\varphi}_1(i), \hat{\lambda}_2\hat{\varphi}_2(i), \dots., \hat{\lambda}_k\hat{\varphi}_k(i)\right] \ i = 1 \dots N. \tag{15}$$

**- Gaussian Granger Causality**

We computed the directed flow between the timeseries in the manifold space by using Granger Causality (Granger, 1980), the quantification of past activity in one location contributing to predict the future activity in another assuming linear dynamics. This corresponds to the unnormalised form of directed transfer entropy under Gaussian assumptions (Deco et al., 2021) inspired by a similar transfer entropy framework from (Brovelli et al., 2015). The approach was developed to infer the underlying bidirectional reciprocal communication characterizing causal interactions between two brain areas by an information theoretical statistical criterion. By using a Gaussian approximation (i.e., only second-order statistics of involved entropies), the method estimates the covariances instead of probabilities, facilitating computation.

Let's assume the following. In order to describe the statistical causal interaction exerted from source brain area *X*, to target brain area *Y*, we measure the extra knowledge that the dynamical functional activity of the past of *X* contributes to the prediction of the future of *Y*. We use the following mutual information:

$$I\left(Y_{i+1}; X^i \middle| Y^i\right) = H\left(Y_{i+1} | Y^i\right) - H\left(Y_{i+1} | X^i, Y^i\right), \tag{16}$$

where $Y_{i+1}$ corresponds to the activity level of brain area *Y* at timepoint *i+1*, $X^i$ is the whole activity level of the past of *X* in a time window of length *T* up to the including timepoint *i* (i.e., $X^i = \left[X_i\ X_{i-1}\ \ldots\ X_{i-(T-1)}\right]$). The time lag between *i* and *i+1* is $t_{lag}$. This causality measure is not symmetric and thus allows bidirectional analysis. Moreover, the conditional entropies are defined as follows:

$$\begin{aligned} H\left(Y_{i+1} | Y^i\right) &= H\left(Y_{i+1}, Y^i\right) - H\left(Y^i\right) \\ &= -\sum_{y_{i+1}, y^i} p\left(y_{i+1}, y^i\right) log\left(p\left(y_{i+1} | y^i\right)\right) \end{aligned} \tag{17}$$

$$\begin{aligned} H\left(Y_{i+1} | X^i, Y^i\right) &= H\left(Y_{i+1}, Y^i, X^i\right) - H\left(X^i, Y^i\right) \\ &= -\sum_{y_{i+1}, y^i, x^i} p\left(y_{i+1}, y^i, x^i\right) log\left(p\left(y_{i+1} | x^i, y^i\right)\right) \end{aligned} \tag{18}$$

The mutual information $I\left(Y_{i+1}; X^i \middle| Y^i\right)$ expresses the degree of statistical dependence between the past of *X* and the future of *Y*. If the mutual information equals zero, the probability $p\left(Y_{i+1}, X^i | Y^i\right) = p\left(Y_{i+1} | Y^i\right). p\left(X^i | Y^i\right)$ and there is no causal interaction from *X* to *Y*.

In consequence, $I\left(Y_{i+1}; X^i \middle| Y^i\right)$ expresses a strong form of Granger Causality (Granger, 1980) by comparing the uncertainty in $Y_{i+1}$ using knowledge of its own past only (i.e., $Y^i$), or the past of both brain regions (i.e., $X^i, Y^i$). The information-theoretical concept of causality was introduced in neuroscience by (Schreiber, 2000) and is usually referred to as transfer entropy (Vicente et al., 2011). A weaker form of causality was proposed by (Brovelli et al., 2015) in order to facilitate computation, by calculating entropies considering Gaussian approximation (i.e., considering only second-order statistics). The entropies can be computed as follows, and causality is based only on the corresponding covariance matrices:

$$H\left(Y^i\right) = \frac{T}{2}\log(2\pi e) + \frac{1}{2}\log\left(det\left(\Sigma\left(Y^i\right)\right)\right), \tag{19}$$

$$H(Y_{i+1}, Y^i) = \frac{T+1}{2}\log(2\pi e) + \frac{1}{2}\log\left(det(\Sigma(Y_{i+1}, Y^i))\right), \tag{20}$$

$$H(X^i, Y^i) = T\log(2\pi e) + \frac{1}{2}\log\left(det(\Sigma(X^i, Y^i))\right), \tag{21}$$

$$H(Y_{i+1}, Y^i, X^i) = \frac{2T+1}{2}\log(2\pi e) + \frac{1}{2}\log\left(det(\Sigma(Y_{i+1}, Y^i, X^i))\right). \tag{22}$$

Heuristically, we did the following, in the manifold space. First, we extracted the lagged versions for each latent *i:*

$$y(i, j - MaxLag, 1{:}p + 1) = [X(i,j),\ X(i, j-1), \ldots, X(i, j-p)]. \tag{23}$$

Then, we quantified the prediction of timeseries *i* from its own past:

$$I_y = logdet(cov(y)) - logdet\left(cov(y(:,2{:}end))\right). \tag{24}$$

The logdet is the logarithm of the determinant of a matrix (i.e., sum of logarithms of the diagonal elements) after Cholesky or LU factorization. Finally, we calculated how the prediction improves when including information from the rest of latents *k*:

$$I_{y|x,z} = I_y - (logdet(cov(z)) - logdet\left(cov(z(:,2{:}end))\right)), \tag{25}$$

where *z* is a matrix combining the lagged values of *i* and *k*. The Granger Causality from *k* to *i* is quantified as this improvement in prediction $I_{y|x,z}$ and saved in matrix *GC(i,k)*. We used a time lag $t_{lag}$=5.

**- Yeo resting-state networks decomposition**

We compared CHARM with the decomposition method using the classical Yeo resting-state network (RSN) at the source space (Yeo et al., 2011). These are: visual (VIS), somatomotor (SOM), dorsal attention (DAN), salience (SAL), limbic (LIM), central executive ( CEN) and default mode network (DMN). We built a map of size 7 Yeo networks * 90 brain areas, representing the number of voxels of cortical and subcortical brain areas belonging to each resting-state networks in the AAL space. For this, we used the mask of the Yeo RSN, and the mask of the AAL parcellation, both defined in MNI152 space. We normalised the map to obtain the probability distribution of regions for each network. Then, we computed the weighted average of our source space brain dynamics (of size 90 brain areas * timepoints) and reduced its dimensionality into 7 Yeo networks * timepoints.

**- Principal component analysis**

We also compared CHARM with Principal component analysis (PCA), a classical linear dimensionality technique. It transforms data into a coordinate system as follows:

$$Y = V^T X, \tag{26}$$

where $V \epsilon \mathcal{R}^{M\, x\, k}$ is a matrix with columns given by the first $k$ eigenvectors - ordered by decreasing order of their corresponding eigenvalues - of the covariance matrix $XX^T \epsilon \mathcal{R}^{M\, x\, M}$. If $k = M,\ V^T$is a rotation matrix satisfying $|\det(V^T)| = 1$ and $V^T = V^{-1}$.

- **Directedness of the reduced space**

We computed a graph theoretical measure of hierarchical organisation to indicate the degree of directedness space (Deco, 2024) to the Gaussian Granger Causality (GC) matrices from the manifold space.

The GC matrix is a graph *C* with *N* features connected by weighted edges determined by the directed flow between the latents in the manifold space. The in-weight ($d_n^{in}$) and out-weight ($d_n^{out}$) for each node (i.e., latent) *n* is calculated by summing the *m* columns and rows of the matrix, respectively:

$$d_n^{in} = \sum_m C_{nm}. \tag{27}$$

$$d_n^{out} = \sum_m C_{mn}. \tag{28}$$

Then, the total weight of a node $u_n$ is defined as follows:

$$u_n = d_n^{in} + d_n^{out}. \tag{29}$$

The imbalance $v_n$ (i.e., difference between the flow into and out of a node) is given by

$$v_n = d_n^{in} + d_n^{out}, \tag{30}$$

and the weighted graph-Laplacian operator $\wedge$ on vector **h** is

$$\wedge = diag(u) - C - C^T. \tag{31}$$

Thus, the following linear system of equations provides the solution **h**:

$$\wedge\, \mathbf{h} = \mathrm{v}. \tag{32}$$

Here, each component of **h** corresponds to the trophic level of each latent. As a note, the operator $\wedge$ is symmetric and the asymmetry comes from the imbalance vector v.

The global directedness is:

$$F_0 = 1 - \frac{\sum_{mn} C_{mn}(h_n - h_m - 1)^2}{\sum_{mn} C_{mn}}. \tag{33}$$

When $F_0 = 1$ there is maximally coherence whilst when $F_0 = 0$ it is considered incoherence.

- **Time asymmetry between networks in manifold space**

We computed the shifted network functional connectivity (FC) between latents in the manifold space by first z-scoring the timeseries across latents, and then measuring the forward time-shifted correlation as follows:

$$FC_{ml}^{shifted} = \frac{\mathfrak{E}[(n_m - \mu_m)(\widehat{\boldsymbol{n}_l} - \mu_l)]}{\sigma_m \sigma_l}. \tag{34}$$

Here, $n_m$ is the timeseries of the latent $m$, and $\widehat{\boldsymbol{n}_l}$ is the timeseries of latent *l* shifted 2 TR forward in time. Moreover, $\mu_m$ and $\mu_l$ are their corresponding mean values across time, and $\sigma_m$ and $\sigma_l$ their standard deviations across time. The expected value operator is $\mathfrak{E}[\,]$.

**- Classification with support vector machine**

We implemented a support vector machine (SVM) for pattern separation of the groups in each CHARM analysis. We used Matlab function *fitcecoc* with a Radial Basis Function (RBF) kernel. The input was the FCtau for each participant with their corresponding unique class label (i.e., group to which they belong to, such as before or after MBCT+TAU during rumination). In particular, the cells of the matrices which were significant when computing statistics between groups. The output was two classes corresponding to the groups compared. We trained the SVM using a leave-one-out cross-validation procedure: one patient was excluded randomly, using the rest for training, testing on the excluded patient, and this was repeated and shuffled 1000 times. The classification was implemented to probe the discriminative nature of the manifolds, specifically whether the significant interactions between latents carry information about treatment changes, rather than as a standalone predictive classifier.

**- Projection of brain dynamics from the source space into each latent**

In order to understand the relation between each brain network and the regional dynamics, we built a functional projection map containing the contribution of the brain dynamics of all brain areas from the source space to each latent of the manifold space. First, we standardized (i.e., z-score) the timeseries of each brain region and latent separately. Then, we calculated the correlation between the timeseries of each brain region and each latent. Specifically, we calculated the Spearman correlation between the source matrix (size timepoints * brain areas), with the manifold matrix (size timepoints * latent dimensions), building a projection map of size brain areas * latent dimensions. Lastly, each column (i.e., latent) of the projection map was normalised by its Euclidean norm. We computed a projection map for the full manifold space and for each group separately, by considering the corresponding subjects' timeseries in the source and reduced space.

**- Comparison of each latent network to reference functional networks**

We evaluated the spatial similarity of the functional projection map of each latent with the RSN (Yeo et al., 2011). To do so, we used the same map from the Yeo RSN decomposition analysis (matrix of size networks * areas with the probability distribution of cortical and subcortical regions for each network). Finally, we computed Pearson's correlation and associated p-values between each resting-state network and each latents functional projection map.

**- Clustering of the manifold space**

CHARM uncovers brain networks in the time domain, therefore we looked at their dynamic reorganisation across groups and relevance to treatment effects. After reducing the dimensionality with CHARM from the source space (90 brain areas * timepoints) to the manifold space (7 latents * timepoints), we looked at what kind of combinations of the latent patterns the brain falls over time. To do so, inspired by the LEiDA method (Cabral et al., 2017), we z-scored and clustered the observables (i.e., timepoints of all subjects), each observable with a number of features equivalent to all latents in the reduced space. We partitioned the manifold space using the K-means clustering, an unsupervised algorithm that assigns data to a cluster centroid to which it has a minimum distance, and iteratively re-calculates the centroids until convergence. The number of clusters used was from $c$=2 to $c$=10, in which each centroid represents a manifold metastate with the dominant latent co-activation (i.e., fingerprint of network configuration). For each clustering result, we calculated the probability of occurrence of each manifold metastate for each subject in each group, by dividing the number of timepoints assigned to a given metastate divided by the total number of timepoints in the scanning session of the subject. Then, we assessed statistically the differences in probabilities of occurrence between groups, and their association with clinical and behavioural scores.

We then obtained the transition probability matrix for each session, and calculated the entropy by modelling transitions as a Markov chain formulation. The entropy rate S is defined as:

$$S = \sum_{i=1}^{N} S_i, \tag{35}$$

where

$$S_i = -p(i) \sum_{j=1}^{N} P(i,j) \log\, P(i,j). \tag{36}$$

Here, $P(i,j)$ represents the transition probability from state $i$ to state $j$, and $p(i)$ denotes the stationary probability of state $i$,. For long realizations of the Markov chain, the probabilities of each state converge to the stationary distribution $p$, solution of

$$P^T p = p. \tag{37}$$

**- Reconstruction in the source space**

In order to assess the robustness of CHARM, we measured the ability of the manifold obtained with CHARM to reconstruct the source data. This is done by first defining a diffusion map on a training set. Then given the $N_G$ timepoints of BOLD signal in $M$ brain areas not included in the definition of the manifold space, we define the generalisation set by $G = [x_1, x_2, \ldots, x_{N_G}] \epsilon\, \mathcal{R}^{M \times N_G}$. The Nyström extension methodology is adopted, derived from the solution of the Fredholm integral equation of the second kind (Evangelou, 2023; Papaioannou et al., 2022; Patsatzis, 2023) described by

$$X_G = X \Psi_k \Lambda_t^{-T} \Psi_k^T P_{G.} \tag{38}$$

Here, $\Psi_k = [\varphi_1, \varphi_2, \dots, \varphi_k]$ is the matrix of the first $k$ eigenvectors of the manifold space. Furthermore, $\Lambda_t$ is the diagonal matrix for the $k$ first eigenvalues at $t$-time diffused steps, whose diagonal elements are $[\lambda_0^t, \lambda_1^t, \dots, \lambda_k^t]$. In the particular case of CHARM, it is the diagonal matrix for the $k$ first eigenvalues of the transition matrix $\hat{P}(t)$ with diagonal elements $[\hat{\lambda}_0, \hat{\lambda}_1, \dots, \hat{\lambda}_k]$. The matrix $P_G \in \mathcal{R}^{N \times N_G}$ is given by the first $N$ rows and last $N_G$ columns of the matrix $\mathrm{P}_{All} \in \mathcal{R}^{(N+N_G)\times(N+N_G)}$ defined by the $t$-time diffused steps transition matrix for all data points (i.e., training and generalisation set). The matrix $\mathrm{P}_{All}$ is computed as $\widehat{W}^t \in \mathcal{R}^{(N+N_G)\times(N+N_G)}$.

To assess the quality of the manifold reduction for each participant, the empirical and reconstructed functional connectivity (FC) are compared.

Given $X$, the matrix BOLD signal observation, timeseries of observations $s_m$ for each brain region $m$ are defined by the $m$-row of $X$. The empirical FC ($FC^{emp}$) is calculated with the Pearson correlation:

$$FC_{ml}^{emp} = \frac{\mathfrak{E}[(s_m-\mu_m)(s_l-\mu_l)]}{\sigma_m\sigma_l}, \tag{39}$$

where for brain areas $m$ and $l$, $\mu_m$ and $\mu_l$ are the mean values across time, respectively, and $\sigma_m$ and $\sigma_l$ are the corresponding standard deviations across time. Moreover, $\mathfrak{E}[]$ is the expected value operator. In addition, the reconstructed FC ($FC^{rec}$) is computed as follows:

$$FC_{ml}^{rec} = \frac{\mathfrak{E}[(r_m-\mu_m)(r_l-\mu_l)]}{\sigma_m\sigma_l}, \tag{40}$$

where $r_m$ are the reconstructed timeseries for the $m$-row of $X$.

To compare $FC^{emp}$ and $FC^{rec}$, their corresponding matrix elements as $f^{emp}$ and $f^{rec}$ are defined. Then, the mean-squared error for $M$ regions is calculated as

$$Err_{FC} = \frac{1}{M^2}\sum_k \left(f_k^{emp} - f_k^{rec}\right)^2 = \frac{1}{M^2}\sum_{m,l}\left(FC_{ml}^{emp} - FC_{ml}^{rec}\right)^2, \tag{41}$$

and the correlation between their elements as

$$Corr_{FC} = \frac{\mathfrak{E}[(f^{emp}-\mu_{emp})(f^{rec}-\mu_{rec})]}{\sigma_{emp}\sigma_{rec}}. \tag{42}$$

Here, $\mu_{emp}$ and $\mu_{rec}$ are the mean values, and $\sigma_{emp}$ and $\sigma_{rec}$ are the standard deviations of the corresponding elements.

**- Inside-out framework**

We compared the causal relationship between the forward pairwise timeseries and their reversed backward version using the INSIDEOUT framework (Deco et al., 2022) in the manifold space.

We consider timeseries $x(t)$ and $y(t)$ evolving from an initial state $A_1$ and $B_1$, respectively, to a final state $A_2$ and $B_2$, respectively. Their reversed backward versions $x^{(r)}(t)$ and $y^{(r)}(t)$ can be obtained by flipping the ordering in time (from $A_2/B_2$ to $A_1/B_1$). The causal dependency between $x(t)$ and $y(t)$ can then be calculated with their time-shifted correlation (at time shift Δt=T, we used T=2) for the forward evolution:

$$c_{forward}(\Delta \mathrm{t}) =< x(t), y(t+\Delta \mathrm{t}) >, \quad (43)$$

and the reversed backward evolution:

$$c_{reversal}(\Delta \mathrm{t}) =< x^{(r)}(t), y^{(r)}(t+\Delta \mathrm{t}). \quad (44)$$

We extended these for the multidimensional case, where the forward and backward evolution of the variable describing the system can be denoted as $x_i(t)$ and $x_i^{(r)}(t)$, and the sub-index $i$ corresponds to the dimensions of the system:

$$FS_{forward,ij}\ (\Delta \mathrm{t}) =< x_i(t), x_j(t+\Delta \mathrm{t}) >, \quad (45)$$

$$FS_{reversal,ij}(\Delta \mathrm{t}) =< x_i{}^{(r)}(t), x_j{}^{(r)}(t+\Delta \mathrm{t}) >. \quad (46)$$

Then we defined a difference matrix $FS_{diff}$, whose elements are the difference of each pair of elements of the forward and reversal matrices:

$$FS_{diff}ij = FS_{forward}ij(\mathrm{T}) - FS_{reversal}ij(\mathrm{T}). \quad (47)$$

Lastly, calculated the irreversibility $I$ of each latent as the mean value elements of $FS_{diff}$ across columns.

### Global brain connectivity

For each subject, we selected the corresponding timepoints in the reduced space and standardized the timeseries (i.e., z-scored). Then, we calculated the Pearson correlation of that matrix and subtracted the identity matrix to exclude self-correlations. Lastly, we calculated the mean to get one value per latent, named global brain connectivity (GBC).

**Figures**

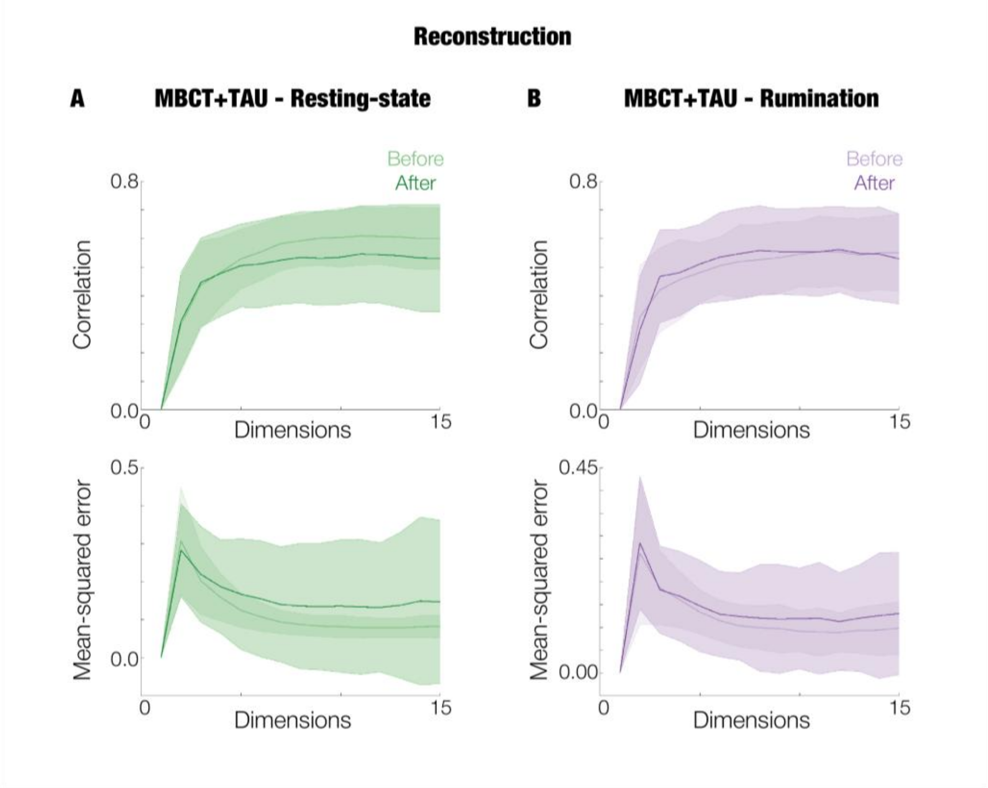


***Supplementary Figure S1. Reconstruction efficacy for CHARM.*** *Fitting (i.e., correlation) in the top row and mean-squared error in bottom the row for* ***A.*** *resting-state MBCT+TAU and* ***B.*** *rumination MBCT+TAU. The mean and standard error of the mean of before and after treatment compared in each analysis are plotted across latents 2-15.*

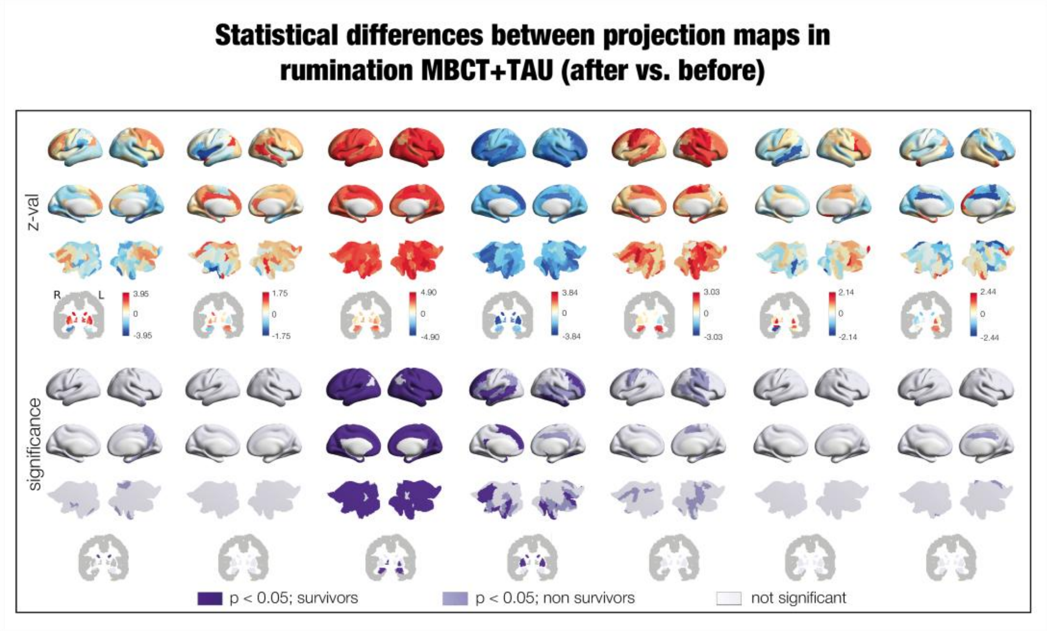


***Supplementary Figure S2. Statistical comparison between the projection maps before and after treatment for rumination MBCT+TAU****. The first row shows the z-value, with positive values (in red) for increases after treatment and the opposite in blue. The second row shows the significance of the comparison, in dark violet and lilac significant brain areas ($p < 0.05$), the former surviving correction for multiple comparisons and the latter not, and in white the regions without significance.*

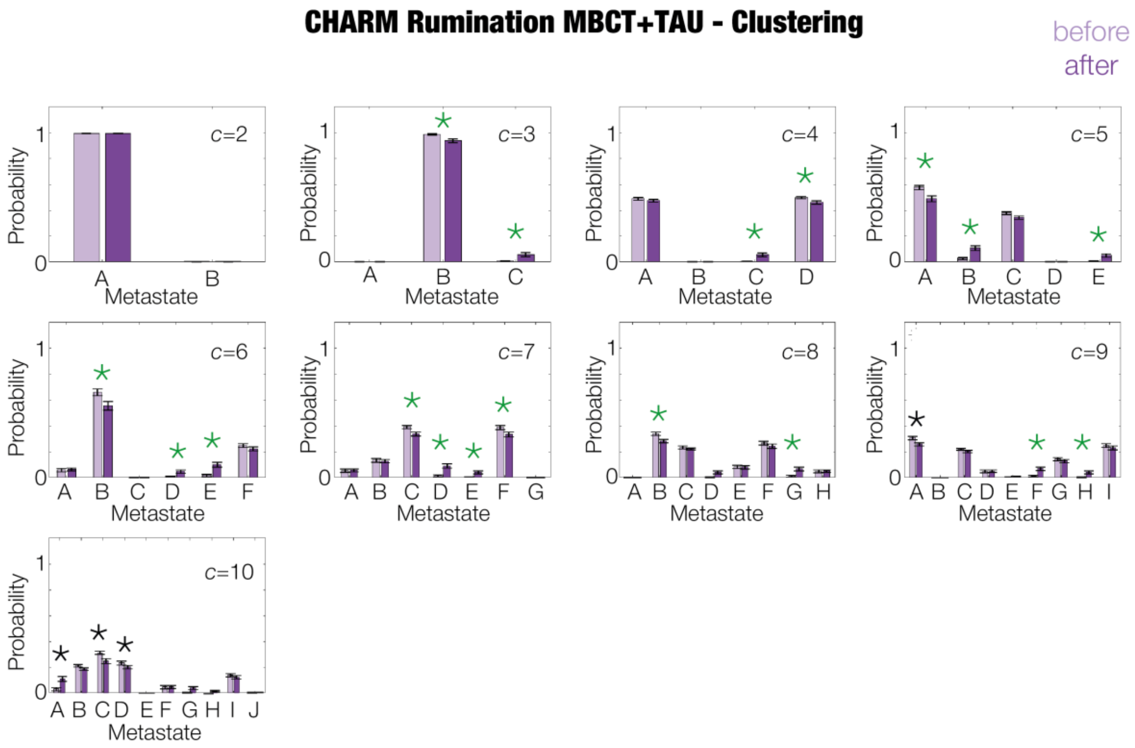


***Supplementary Figure S3.*** *Differences between probabilities of occurrence of metastates across different number of cluster centres (k=2-10).*

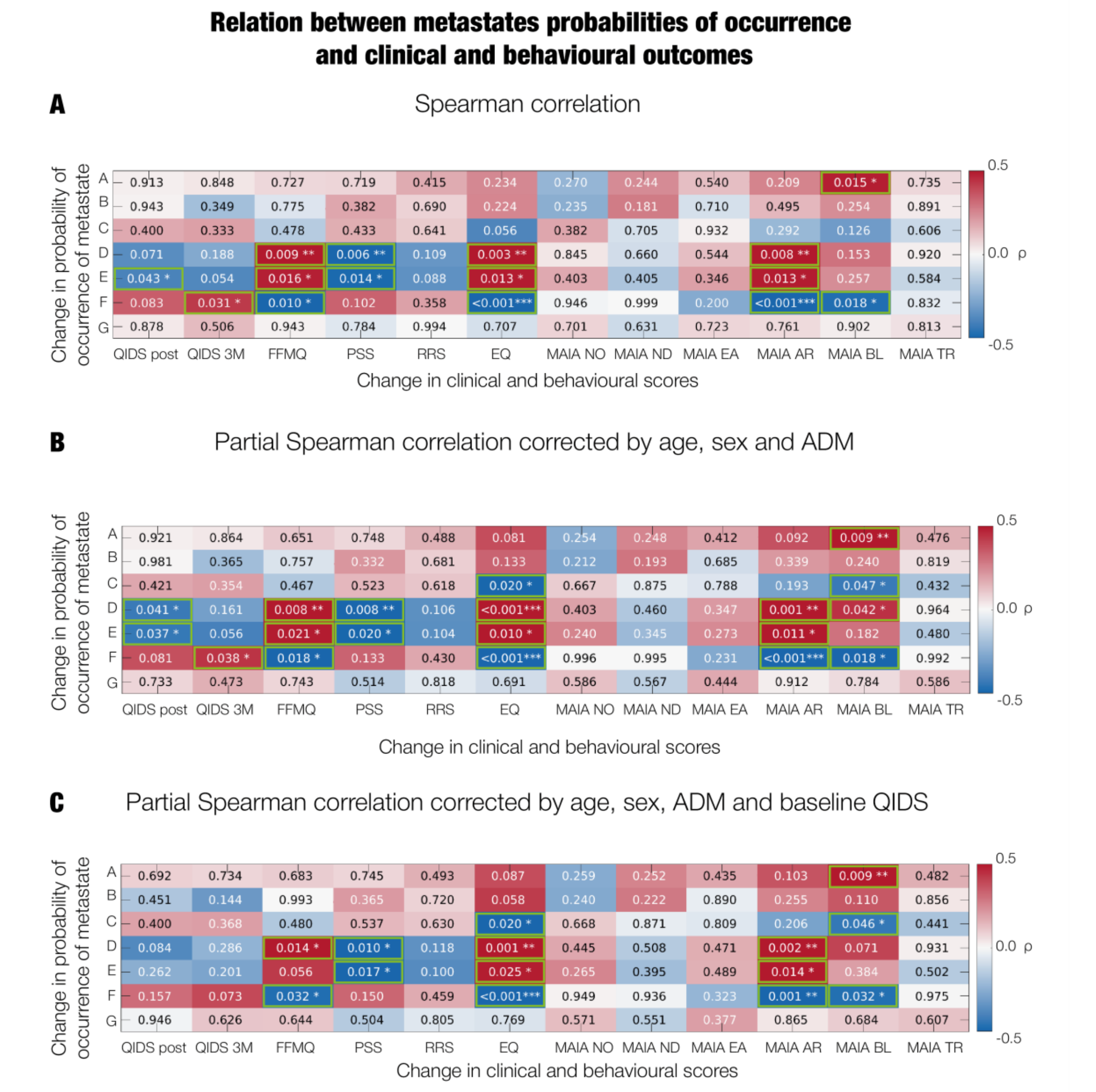


***Supplementary Figure S4. Relation between clustering results and behavioural and clinical and behavioural scores.*** *Correlation between change in probability of occurrence of each metastate, and change in clinical and behavioural scores.* ***A.*** *Spearman correlation* ***B.*** *Partial Spearman correlation corrected by covariates of age, sex and ADM.* ***C.*** *Partial Spearman correlation corrected by covariates of age, sex and ADM as well as baseline QIDS. The strength of each correlation can be identified in the colourbar. The significance is written in each cell, with asterisks representing the significant differences (*, p < 0.05; **, p < 0.01; ***, p < 0.001). No correction for multiple comparisons was done for either case given this analysis is exploratory*

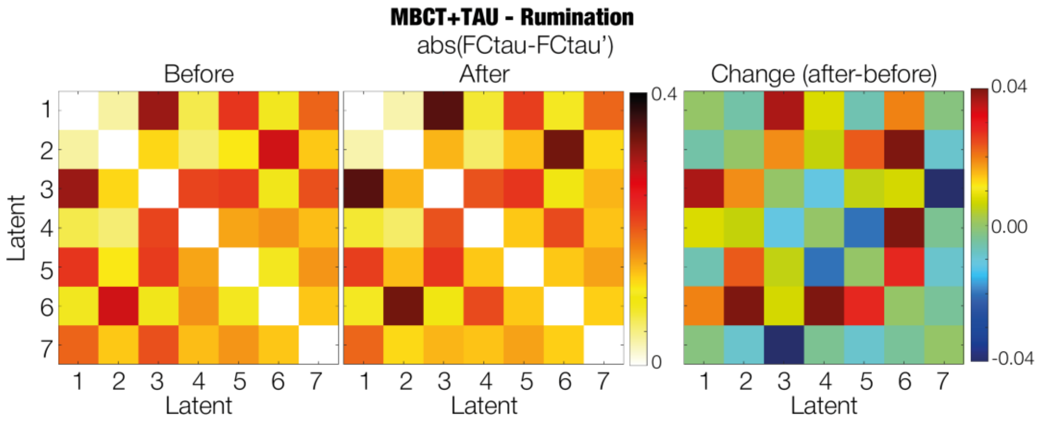


***Supplementary Figure S5. Time asymmetries in rumination MBCT+TAU obtained with CHARM.*** *Left matrices show the absolute difference of the shifted functional connectivity subtracted with its transpose for before and after treatment. The strength is represented by the colourbar (red corresponding to higher values). The right matrix shows their difference, with positive values in red and negative in blue.*

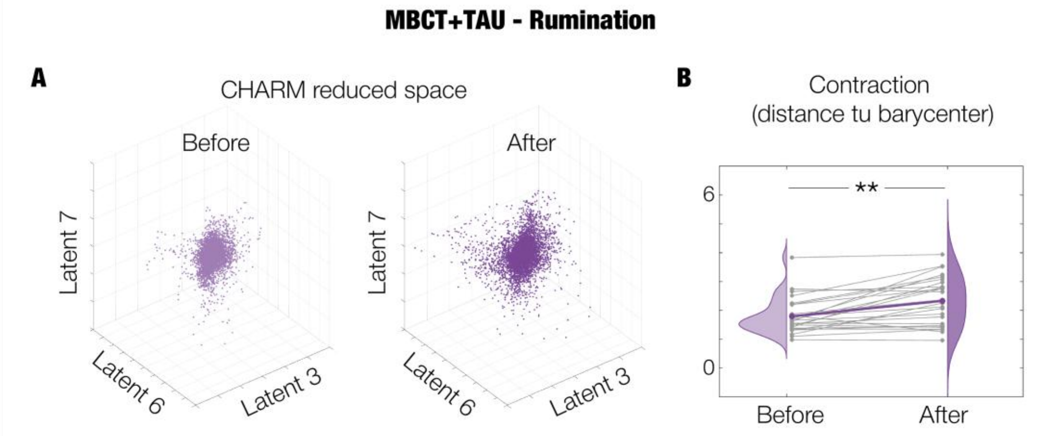


***Supplementary Figure S6. Dispersion of CHARM latent space in rumination MBCT+TAU. A.*** *The 3D scatterplots show timepoints distributed along 3 gradients for before and after treatment.* ***B.*** *We obtained the cloud centre (i.e., braycentre) of the 7-dimensional manifold space and calculated the distance of each timepoint to this barycentre. Grey lines represent the trajectory of each individual whereas the coloured line represents their averaged trajectory. This distance significantly increases after treatment (**, $p < 0.01$).*

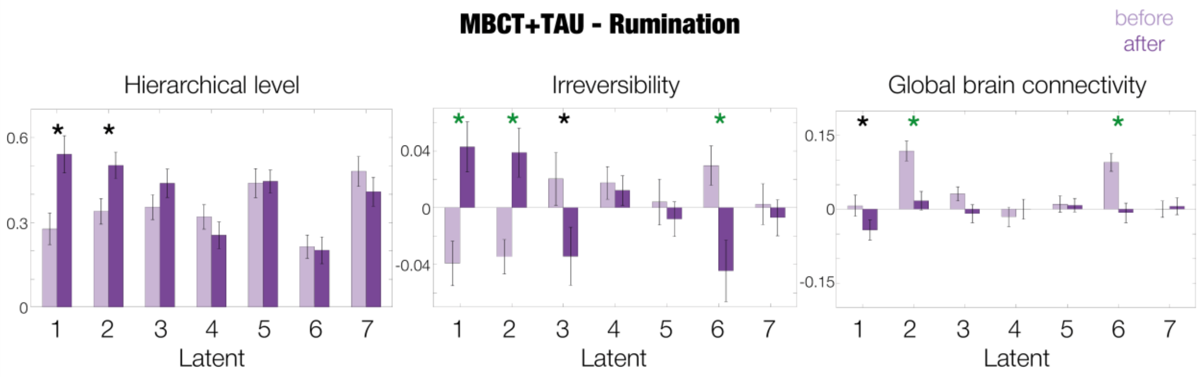


***Supplementary Figure S7. Brain measures of latent networks in rumination MBCT+TAU.*** *Barplots for the hierarchical level, irreversibility, and global brain connectivity of before and after treatment for all latents in the reduced manifold space. Significance is represented by an asterisk (*, $p < 0.05$), in green the survivors after correction for multiple comparisons within each analysis.*

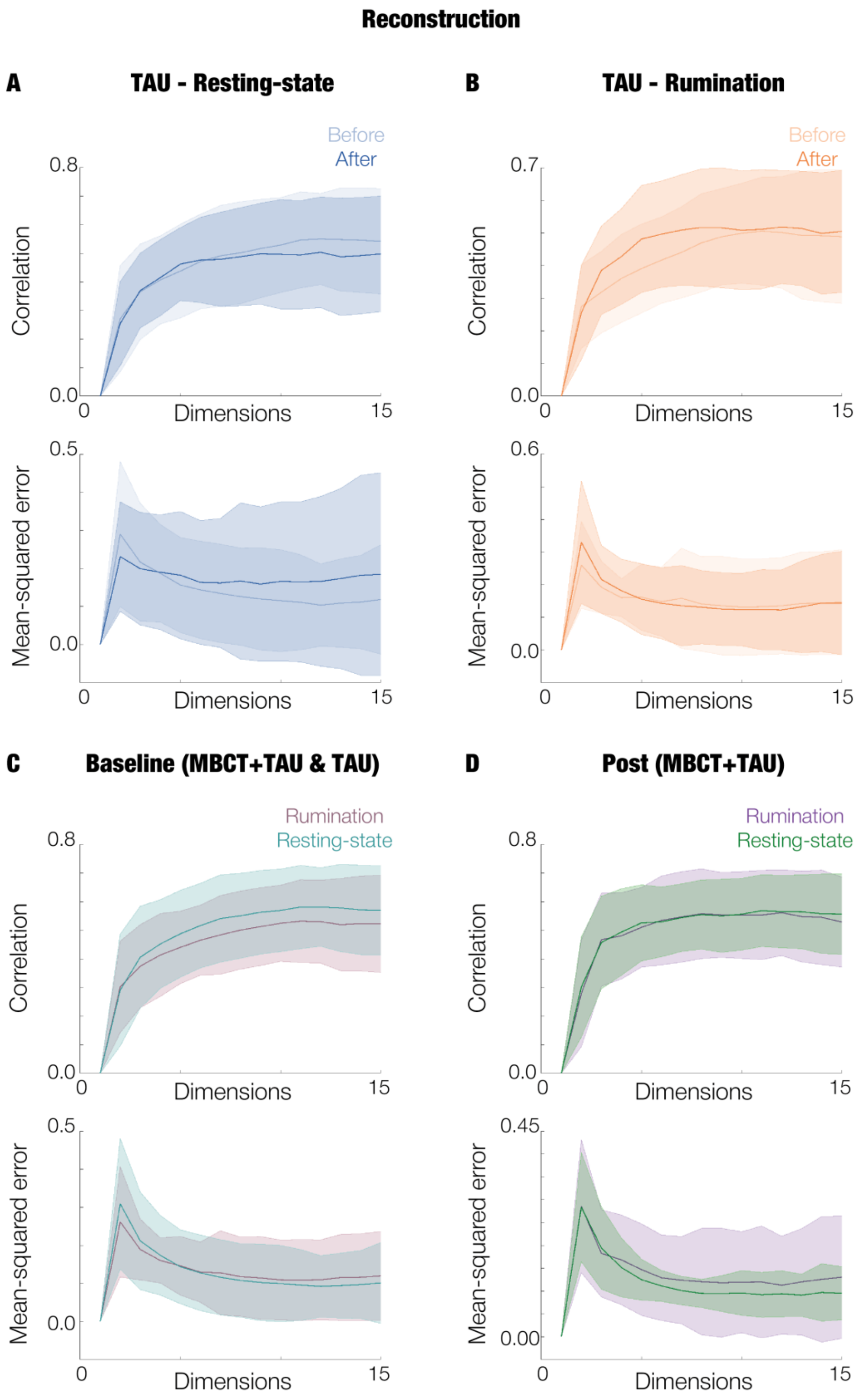


***Supplementary Figure 8. Reconstruction efficacy for CHARM.*** *Fitting (i.e., correlation) and mean-squared error across latents 2-15 for* ***A.*** *resting-state TAU,* ***B.*** *rumination TAU,* ***C.*** *baseline MBCT+TAU and TAU and* ***D.*** *post-treatment MBCT+TAU. The lines represent the mean and the shades represent the standard error of the mean of the groups.*

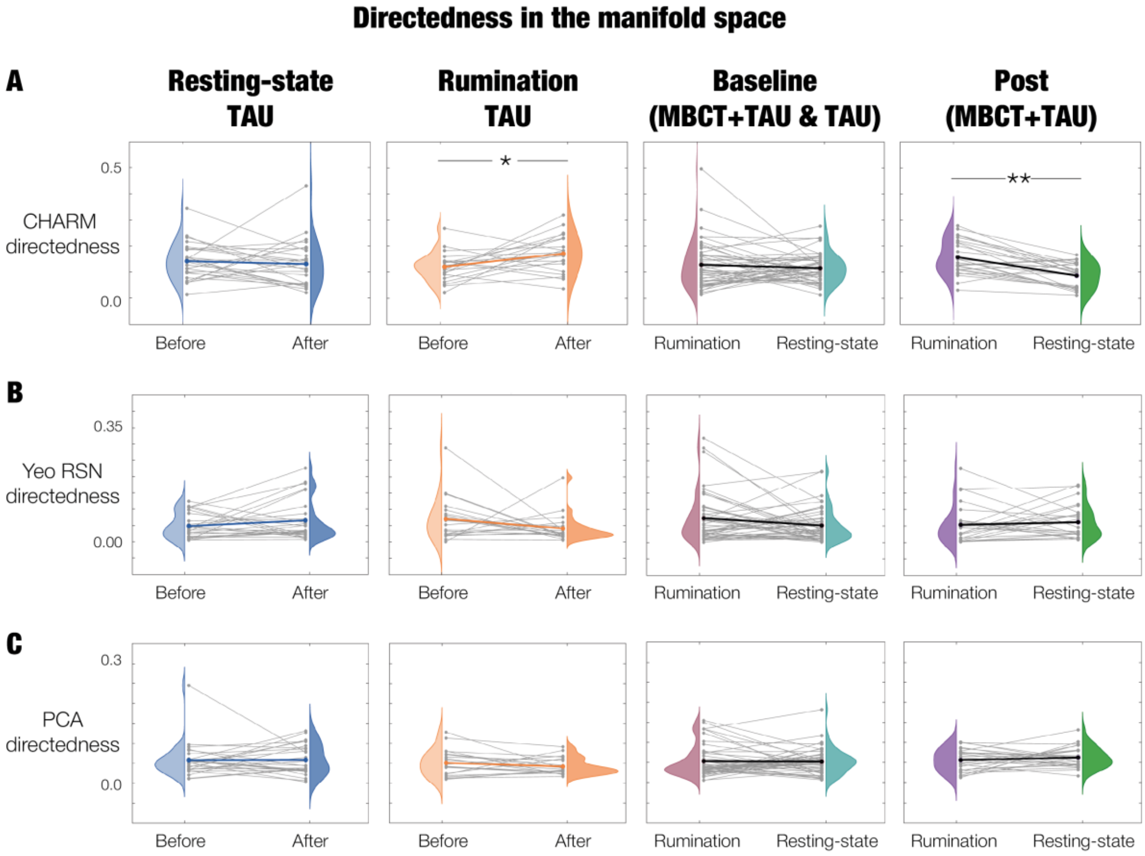


***Supplementary Figure S9. Global directedness in latent space for the rest of the treatments and scan conditions.*** *Changes in brain hierarchy calculated on the Granger Causality matrices in the reduced space following* ***A.*** *CHARM* ***B.*** *Yeo networks and* ***C.*** *PCA. Significance is represented by asterisks (*, p < 0.05; **, p < 0.01). Grey lines represent the trajectory of each individual whereas the coloured line represents their averaged trajectory.*

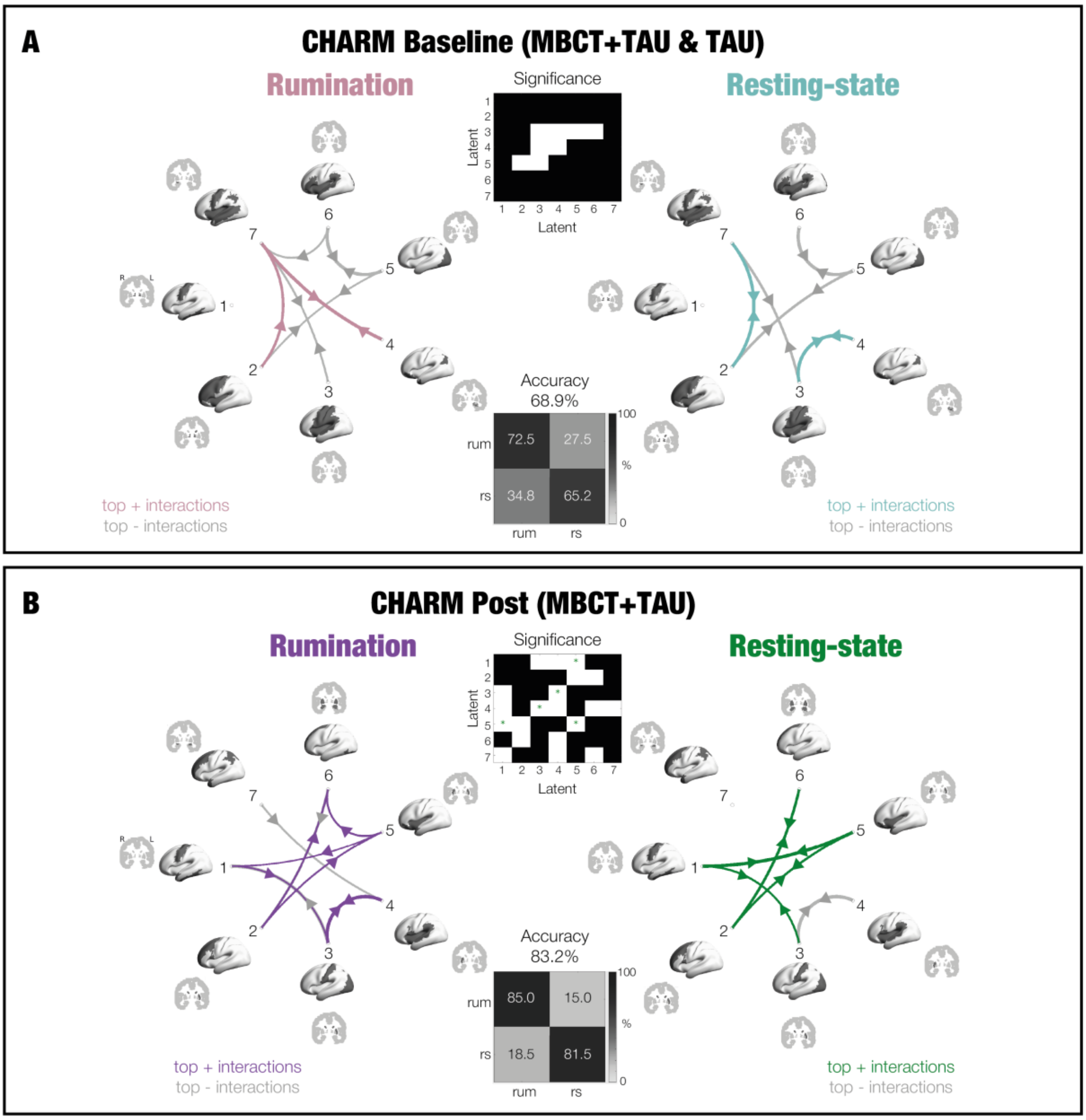


***Supplementary Figure S10. Top network interactions at baseline and post-treatment captured with CHARM.*** *Differences between resting-state and rumination treatment at* ***A.*** *baseline, combining MBCT+TAU and TAU, and* ***B.*** *after treatment, for the MBCT+TAU group. Circular maps show top 20% of interactions obtained from the shifted network functional connectivity matrices, with strongest positive and negative interactions, and arrow width representing their strength. The black and white matrices show the significance between scan condition (resting-state and rumination) in white ($p < 0.05$), and survivors after correction by multiple comparisons are identified with a green asterisk. Importantly, the schematics depicting top 20% of latent interactions within each group align partially with the significant differences identified by the white squares in the matrices, as these reflect different comparisons (the former shows the within-group asymmetric interactions whereas the latter shows the between group statistical differences between these interactions). Confusion matrices show the classification of significant interactions using a support vector machine.*

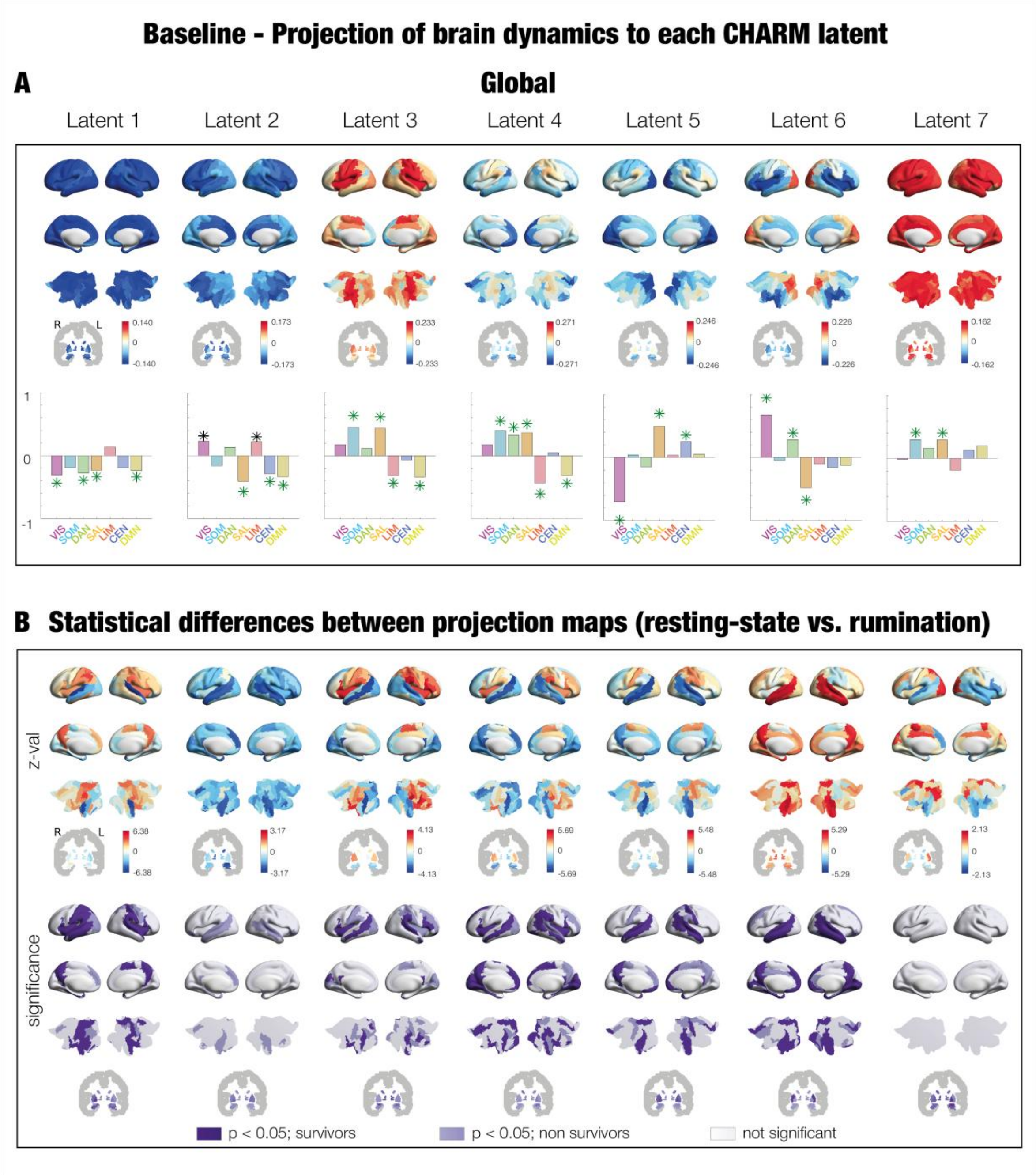


***Supplementary Figure S11. Baseline projection of brain dynamics into each manifold network. A.*** *For the CHARM manifold space, the brain renders show the contribution of the whole-brain dynamics in the source space to each latent of the reduced space (i.e., projection map). We also calculated the correlation between these contributions and the prototypical Yeo RSN.* ***B.*** *Statistical comparison between the projection maps of rumination and resting-state. The first row shows the z-value, with positive values (in red) for higher values in resting-state compared to rumination, and the opposite in blue. The second row shows the significance of the comparison, in dark violet and lilac significant brain areas ($p < 0.05$), the former surviving correction for multiple comparisons and the latter not, and in white the regions without significance. Subcortical areas are shown on slices in Montreal Neurological Institute (MNI) space (coronal axis y=-6 mm).*

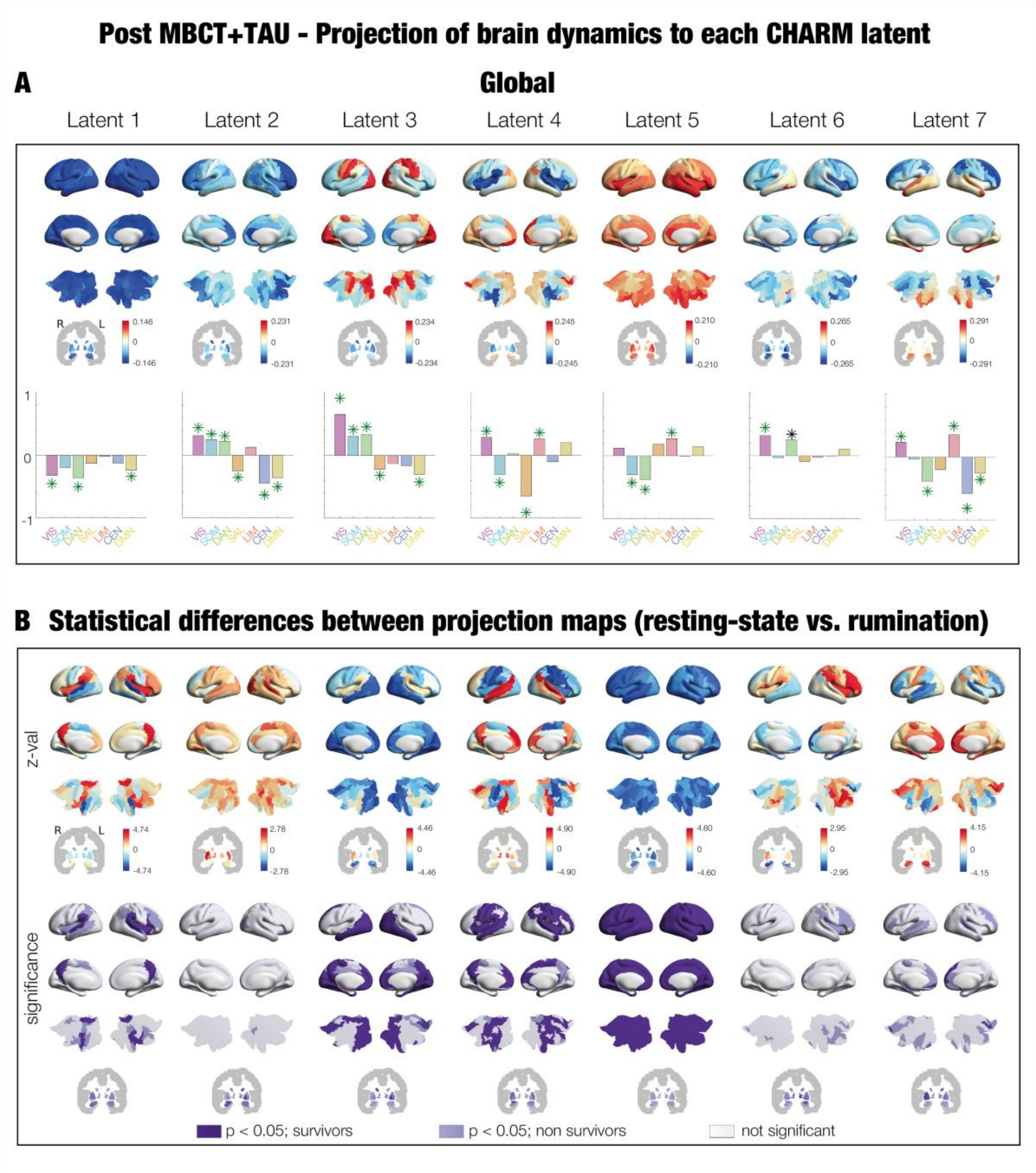


***Supplementary Figure S12. Post MBCT+TAU projection of brain dynamics into each manifold network. A.*** *For the CHARM manifold space, the brain renders show the contribution of the whole-brain dynamics in the source space to each latent of the reduced space (i.e., projection map). We also calculated the correlation between these contributions and the prototypical Yeo RSN.* ***B.*** *Statistical comparison between the projection maps of rumination and resting-state. The first row shows the z-value, with positive values (in red) for higher values in resting-state compared to rumination, and the opposite in blue. The second row shows the significance of the comparison, in dark violet and lilac significant brain areas (p < 0.05), the former surviving correction for multiple comparisons and the latter not, and in white the regions without significance. Subcortical areas are shown on slices in Montreal Neurological Institute (MNI) space (coronal axis y=-6 mm).*

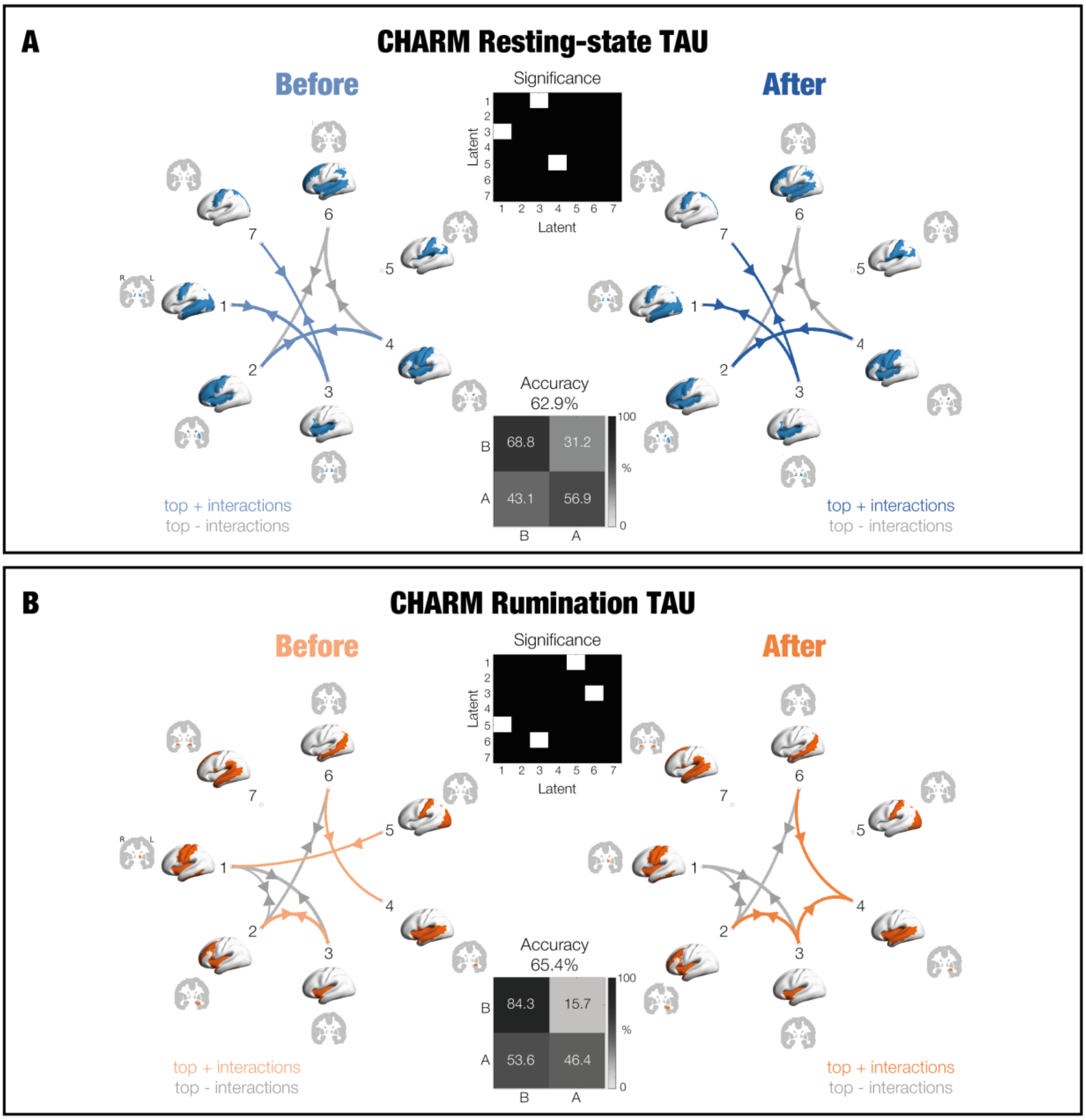


***Supplementary Figure S13. Top network interaction in TAU captured with CHARM.*** *Differences between before and after treatment in **A.** resting-state and **B.** rumination. Time asymmetries are schematised in the circular maps, with strongest positive and negative interactions, and arrow width representing their strength. Significance between sessions (before and after) is represented in the white squares of the black and white matrices (p < 0.05). Importantly, the schematics depicting top 20% of latent interactions within each group align partially with the significant differences identified by the white squares in the matrices, as these reflect different comparisons (the former shows the within-group asymmetric interactions whereas the latter shows the between group statistical differences between these interactions). Confusion matrices show the classification of significant interactions using a support vector machine.*

**Tables**

| RUM MBCT+TAU change (after-before) | Spearman | | Partial Spearman corrected by age, sex & mADM | | Partial Spearman corrected by age, sex, mADM & QIDS0 | | number of subjects (N) |
|---|---|---|---|---|---|---|---|
| | Rho | P-val | Rho | P-val | Rho | P-val | |
| **Mental health outcomes** | | | | | | | |
| **QIDS POST** | **-0.391** | **0.048** | **-0.446** | **0.033** | -0.287 | 0.196 | 26 |
| **QIDS 3M** | -0.234 | 0.249 | -0.253 | 0.245 | -0.116 | 0.606 | 26 |
| **PSS** | -0.242 | 0.243 | -0.324 | 0.141 | -0.325 | 0.151 | 25 |
| **Mechanism outcomes** | | | | | | | |
| **FFMQ_TOTAL** | 0.350 | 0.086 | 0.354 | 0.106 | 0.277 | 0.225 | 25 |
| **RRS_Total** | -0.181 | 0.387 | -0.166 | 0.459 | -0.159 | 0.490 | 25 |
| **EQ** | **0.524** | **0.007** | **0.441** | **0.040** | 0.386 | 0.084 | 25 |
| **MAIA_NO** | 0.314 | 0.127 | 0.197 | 0.379 | 0.183 | 0.427 | 25 |
| **MAIA_ND** | -0.177 | 0.396 | -0.137 | 0.544 | -0.115 | 0.619 | 25 |
| **MAIA_EA** | **0.436** | **0.029** | 0.381 | 0.080 | 0.321 | 0.156 | 25 |
| **MAIA_AR** | **0.495** | **0.012** | 0.401 | 0.065 | 0.378 | 0.091 | 25 |
| **MAIA_BL** | **0.410** | **0.042** | 0.359 | 0.101 | 0.282 | 0.216 | 25 |
| **MAIA_TR** | -0.013 | 0.950 | -0.191 | 0.395 | -0.188 | 0.414 | 25 |

***Table S1: Correlations between change (post-pre) scores in directedness of the manifold space with change (post-pre) in psychological processes and clinical outcomes for MBCT+TAU rumination.*** *Columns: For each correlation (Spearman or Partial Spearman) we report the strength, significance, and number of subjects with response in all variables involved (N). Significant correlations are in bold and green. Rows abbreviations: QIDS post treatment and 3 months follow up; PSS, perceived stress; FFMQ, Five Factor Mindfulness Questionnaire; RRS, rumination response scale; EQ, Experience Questionnaire; MAIA, Multidimensional Assessment of Interoceptive Awareness subscores of NO, noticing, ND, not-distracting, EA, emotional awareness, AR, attention regulation, BL, body listening, and TR, trusting.*

| RS1 MBCT+TAU change (after-before) | Spearman | | Partial Spearman corrected by age, sex & mADM | | Partial Spearman corrected by age, sex, mADM & QIDS0 | | number of subjects (N) |
|---|---|---|---|---|---|---|---|
| | Rho | P-val | Rho | P-val | Rho | P-val | |
| **Mental health outcomes** | | | | | | | |
| **QIDS POST** | -0.010 | 0.952 | 0.059 | 0.731 | 0.174 | 0.319 | 39 |
| **QIDS 3M** | 0.000 | 0.998 | 0.009 | 0.959 | 0.068 | 0.702 | 38 |
| **PSS** | 0.027 | 0.872 | 0.059 | 0.739 | 0.083 | 0.645 | 37 |
| **Mechanism outcomes** | | | | | | | |
| **FFMQ_TOTAL** | -0.028 | 0.869 | -0.006 | 0.971 | -0.027 | 0.879 | 37 |
| **RRS_Total** | 0.168 | 0.319 | 0.171 | 0.333 | 0.192 | 0.285 | 37 |
| **EQ** | -0.231 | 0.168 | -0.154 | 0.385 | -0.189 | 0.291 | 37 |
| **MAIA_NO** | -0.121 | 0.474 | -0.120 | 0.499 | -0.122 | 0.500 | 37 |
| **MAIA_ND** | 0.137 | 0.419 | 0.128 | 0.469 | 0.115 | 0.524 | 37 |
| **MAIA_EA** | 0.110 | 0.518 | 0.100 | 0.572 | 0.078 | 0.665 | 37 |
| **MAIA_AR** | 0.007 | 0.968 | 0.129 | 0.467 | 0.117 | 0.516 | 37 |
| **MAIA_BL** | 0.026 | 0.879 | 0.020 | 0.911 | -0.023 | 0.901 | 37 |
| **MAIA_TR** | 0.123 | 0.467 | 0.194 | 0.271 | 0.173 | 0.337 | 37 |

***Table S2: Correlations between change (post-pre) scores in directedness of the manifold space with change (post-pre) in psychological processes and clinical outcomes for MBCT+TAU resting-state.*** *Columns: For each correlation (Spearman and Partial Spearman) we report the strength, significance, and number of subjects with response in all variables involved (N). There are no significant correlations. Rows abbreviations: QIDS post treatment and 3 months follow up; PSS, perceived stress; FFMQ, Five Factor Mindfulness Questionnaire; RRS, rumination response scale; EQ, Experience Questionnaire; MAIA, Multidimensional Assessment of Interoceptive Awareness subscores of NO, noticing, ND, not-distracting, EA, emotional awareness, AR, attention regulation, BL, body listening, and TR, trusting.*

| | Top positive | | Bottom negative | |
|---|---|---|---|---|
| 1 | - | | 61. Parietal_Inf_L | -0.116 |
| | | | 8. Frontal_Mid_R | -0.117 |
| | | | 1. Precentral_L | -0.118 |
| | | | 3. Frontal_Sup_L | -0.118 |
| | | | 30. Insula_R | -0.118 |
| | | | 29, Insula_L | -0.119 |
| | | | 89. Temporal_Inf_L | -0.120 |
| | | | 4. Frontal_Sup_R | -0.120 |
| | | | 56. Fusiform_R | -0.121 |
| | | | 19. Supp_Motor_Area_L | -0.122 |
| | | | 55. Fusiform_L | -0.124 |
| | | | 20. Supp_Motor_Area_R | -0.125 |
| | | | 68. Precuneus_R | -0.125 |
| | | | 90. Temporal_Inf_R | -0.125 |
| | | | 86. Temporal_Mid_R | -0.126 |
| | | | 67. Precuneus_L | -0.128 |
| | | | 34. Cingulum_Mid_R | -0.130 |
| | | | 33. Cingulum_Mid_L | -0.131 |
| 2 | 72. Caudate_R | 0.181 | - | |
| | 14. Frontal_Inf_Tri_R | 0.180 | | |
| | 32. Cingulum_Ant_R | 0.179 | | |
| | 16. Frontal_Inf_Orb_R | 0.177 | | |
| | 24. Frontal_Sup_Medial_R | 0.165 | | |
| | 73. Putamen_L | 0.161 | | |
| | 23. Frontal_Sup_Medial_L | 0.161 | | |
| | 71. Caudate_L | 0.158 | | |
| | 11. Frontal_Inf_Oper_L | 0.154 | | |
| | 86. Temporal_Mid_R | 0.153 | | |
| | 4. Frontal_Sup_R | 0.152 | | |
| | 38. Hippocampus_R | 0.145 | | |
| | 31. Cingulum_Ant_L | 0.143 | | |
| | 8. Frontal_Mid_R | 0.142 | | |
| | 30. Insula_R | 0.139 | | |
| | 3. Frontal_Sup_L | 0.138 | | |
| | 12. Frontal_Inf_Oper_R | 0.136 | | |
| | 65. Angular_L | 0.136 | | |
| 3 | 46. Cuneus_R | 0.232 | 13. Frontal_Inf_Tri_L | -0.089 |
| | 49. Occipital_Sup_L | 0.224 | 9. Frontal_Mid_Orb_L | -0.090 |
| | 45. Cuneus_L | 0.220 | 14. Frontal_Inf_Tri_R | -0.092 |
| | 50. Occipital_Sup_R | 0.217 | 10. Frontal_Mid_Orb_R | -0.092 |
| | 47. Lingual_L | 0.199 | 24. Frontal_Sup_Medial_R | -0.096 |
| | 44. Calcarine_R | 0.198 | 23. Frontal_Sup_Medial_L | -0.097 |
| | 48. Lingual_R | 0.194 | 78. Thalamus_R | -0.102 |
| | 52. Occipital_Mid_R | 0.189 | 15. Frontal_Inf_Orb_L | -0.108 |
| | 43. Calcarine_L | 0.189 | 16. Frontal_Inf_Orb_R | -0.111 |
| | 51. Occipital_Mid_L | 0.180 | 75. Pallidum_L | -0.112 |
| | 58. Postcentral_R | 0.173 | 31. Cingulum_Ant_L | -0.114 |
| | 57. Postcentral_L | 0.163 | 32. Cingulum_Ant_R | -0.116 |

| | | | | |
|---|---|---|---|---|
| | 53. Occipital_Inf_L | 0.153 | 77. Thalamus_L | -0.119 |
| | 2. Precentral_R | 0.152 | 76. Pallidum_R | -0.120 |
| | 69. Paracentral_Lobule_L | 0.148 | 74. Putamen_R | -0.129 |
| | 70. Paracentral_Lobule_R | 0.139 | 71. Caudate_L | -0.145 |
| | 54. Occipital_Inf_R | 0.139 | 73. Putamen_L | -0.148 |
| | 60. Parietal_Sup_R | 0.128 | 72. Caudate_R | -0.155 |
| **4** | | | 39. ParaHippocampal_L | -0.129 |
| | | | 40. ParaHippocampal_R | -0.130 |
| | | | 42. Amygdala_R | -0.131 |
| | | | 56. Fusiform_R | -0.132 |
| | | | 72. Caudate_R | -0.133 |
| | | | 87. Temporal_Pole_Mid_L | -0.133 |
| | | | 84. Temporal_Pole_Sup_R | -0.136 |
| | | | 75. Pallidum_L | -0.137 |
| | | | 14. Frontal_Inf_Tri_R | -0.138 |
| | | | 86. Temporal_Mid_R | -0.142 |
| | | | 38. Hippocampus_R | -0.142 |
| | | | 30. Insula_R | -0.147 |
| | | | 83. Temporal_Pole_Sup_L | -0.149 |
| | | | 88. Temporal_Pole_Mid_R | -0.152 |
| | | | 16. Frontal_Inf_Orb_R | -0.156 |
| | | | 74. Putamen_R | -0.158 |
| | | | 29, Insula_L | -0.162 |
| | | | 73. Putamen_L | -0.165 |
| **5** | 27. Rectus_L | 0.284 | 83. Temporal_Pole_Sup_L | -0.073 |
| | 28. Rectus_R | 0.282 | 75. Pallidum_L | -0.074 |
| | 6. Frontal_Sup_Orb_R | 0.222 | 74. Putamen_R | -0.078 |
| | 21. Olfactory_L | 0.222 | 78. Thalamus_R | -0.079 |
| | 26. Frontal_Med_Orb_R | 0.217 | 76. Pallidum_R | -0.079 |
| | 25. Frontal_Med_Orb_L | 0.214 | 64. SupraMarginal_R | -0.083 |
| | 22. Olfactory_R | 0.213 | 77. Thalamus_L | -0.086 |
| | 5. Frontal_Sup_Orb_L | 0.204 | 29, Insula_L | -0.093 |
| | 10. Frontal_Mid_Orb_R | 0.193 | 85. Temporal_Mid_L | -0.105 |
| | 9. Frontal_Mid_Orb_L | 0.123 | 30. Insula_R | -0.106 |
| | 24. Frontal_Sup_Medial_R | 0.104 | 84. Temporal_Pole_Sup_R | -0.123 |
| | 16. Frontal_Inf_Orb_R | 0.092 | 63. SupraMarginal_L | -0.154 |
| | 66. Angular_R | 0.092 | 18. Rolandic_Oper_R | -0.156 |
| | 32. Cingulum_Ant_R | 0.088 | 17. Rolandic_Oper_L | -0.169 |
| | 23. Frontal_Sup_Medial_L | 0.087 | 82. Temporal_Sup_R | -0.212 |
| | 4. Frontal_Sup_R | 0.081 | 80. Heschl_R | -0.230 |
| | 31. Cingulum_Ant_L | 0.079 | 81. Temporal_Sup_L | -0.236 |
| | 39. ParaHippocampal_L | 0.075 | 79. Heschl_L | -0.241 |
| **6** | 90. Temporal_Inf_R | 0.231 | 51. Occipital_Mid_L | -0.004 |
| | 14. Frontal_Inf_Tri_R | 0.222 | 43. Calcarine_L | -0.007 |
| | 80. Heschl_R | 0.221 | 63. SupraMarginal_L | -0.025 |
| | 10. Frontal_Mid_Orb_R | 0.203 | 59. Parietal_Sup_L | -0.027 |
| | | | 49. Occipital_Sup_L | -0.041 |
| | | | 53. Occipital_Inf_L | -0.121 |

| | | |
|---|---|---|
| | 16. Frontal_Inf_Orb_R 0.192<br>30. Insula_R 0.182<br>4. Frontal_Sup_R 0.176<br>27. Rectus_L 0.176<br>76. Pallidum_R 0.159<br>21. Olfactory_L 0.157<br>9. Frontal_Mid_Orb_L 0.157<br>88. Temporal_Pole_Mid_R 0.149<br>56. Fusiform_R 0.147<br>2. Precentral_R 0.146<br>52. Occipital_Mid_R 0.146<br>6. Frontal_Sup_Orb_R 0.146<br>38. Hippocampus_R 0.141<br>26. Frontal_Med_Orb_R 0.141 | |
| **7** | 62. Parietal_Inf_R 0.254<br>8. Frontal_Mid_R 0.234<br>4. Frontal_Sup_R 0.207<br>60. Parietal_Sup_R 0.195<br>7. Frontal_Mid_L 0.187<br>12. Frontal_Inf_Oper_R 0.171<br>61. Parietal_Inf_L 0.165<br>3. Frontal_Sup_L 0.165<br>66. Angular_R 0.163<br>24. Frontal_Sup_Medial_R 0.154<br>64. SupraMarginal_R 0.153<br>59. Parietal_Sup_L 0.152<br>20. Supp_Motor_Area_R 0.143<br>19. Supp_Motor_Area_L 0.133<br>1. Precentral_L 0.133<br>14. Frontal_Inf_Tri_R 0.131<br>23. Frontal_Sup_Medial_L 0.126<br>32. Cingulum_Ant_R 0.125 | 82. Temporal_Sup_R -0.077<br>85. Temporal_Mid_L -0.077<br>80. Heschl_R -0.087<br>79. Heschl_L -0.094<br>89. Temporal_Inf_L -0.099<br>55. Fusiform_L -0.104<br>81. Temporal_Sup_L -0.106<br>84. Temporal_Pole_Sup_R -0.109<br>56. Fusiform_R -0.113<br>38. Hippocampus_R -0.115<br>37. Hippocampus_L -0.126<br>83. Temporal_Pole_Sup_L -0.133<br>41. Amygdala_L -0.140<br>42. Amygdala_R -0.141<br>88. Temporal_Pole_Mid_R -0.176<br>87. Temporal_Pole_Mid_L -0.183<br>39. ParaHippocampal_L -0.191<br>40. ParaHippocampal_R -0.209 |

***Supplementary Table S3. Rumination MBCT+TAU projection of brain dynamics into each manifold network.*** *We obtained the contribution of the whole-brain dynamics in the source space to each latent of the reduced space by calculating the correlation between the timeseries of each brain region with the timeseries of each latent. The strongest 20% of contribution, positive (left column) and negative (right column) for each latent can be seen in each row.*

| Latent | Area, RSN, tstat, p, survivor | | | |
|---|---|---|---|---|
| 1 | 72. Caudate_R | 3.594 | 0.000325873 | ✓ |
| | 71. Caudate_L | 3.264 | 0.001099908 | ✓ |
| | 73. Putamen_L | 3.060 | 0.00221 | x |
| | 74. Putamen_R | 2.806 | 0.005008657 | x |
| | 40. ParaHippocampal_R | -2.781 | 0.005417863 | x |
| | 77. Thalamus_L | 2.781 | 0.005417863 | x |
| | 78. Thalamus_R | 2.578 | 0.009940429 | x |
| | 38. Hippocampus_R | -2.527 | 0.011500916 | x |
| | 88. Temporal_Pole_Mid_R | -2.375 | 0.017562701 | x |
| | 39. ParaHippocampal_L | -2.324 | 0.020130054 | x |
| | 17. Rolandic_Oper_L | -2.121 | 0.033944438 | x |
| | 75. Pallidum_L | 2.121 | 0.033944438 | x |
| | 68. Precuneus_R | -1.994 | 0.046180343 | x |
| 2 | - | | | |
| 3 | 14. Frontal_Inf_Tri_R | 4.457 | 8.2981E-06 | ✓ |
| | 12. Frontal_Inf_Oper_R | 4.280 | 1.87264E-05 | ✓ |
| | 84. Temporal_Pole_Sup_R | 4.102 | 4.09996E-05 | ✓ |
| | 9. Frontal_Mid_Orb_L | 3.975 | 7.04433E-05 | ✓ |
| | 16. Frontal_Inf_Orb_R | 3.899 | 9.67546E-05 | ✓ |
| | 86. Temporal_Mid_R | 3.899 | 9.67546E-05 | ✓ |
| | 90. Temporal_Inf_R | 3.899 | 9.67546E-05 | ✓ |
| | 83. Temporal_Pole_Sup_L | 3.670 | 0.000242543 | ✓ |
| | 64. SupraMarginal_R | 3.645 | 0.000267798 | ✓ |
| | 89. Temporal_Inf_L | 3.645 | 0.000267798 | ✓ |
| | 20. Supp_Motor_Area_R | 3.619 | 0.000295502 | ✓ |
| | 11. Frontal_Inf_Oper_L | 3.594 | 0.000325873 | ✓ |
| | 87. Temporal_Pole_Mid_L | 3.594 | 0.000325873 | ✓ |
| | 5. Frontal_Sup_Orb_L | 3.568 | 0.000359146 | ✓ |
| | 58. Postcentral_R | 3.543 | 0.000395576 | ✓ |
| | 2. Precentral_R | 3.492 | 0.00047902 | ✓ |
| | 13. Frontal_Inf_Tri_L | 3.492 | 0.00047902 | ✓ |
| | 15. Frontal_Inf_Orb_L | 3.492 | 0.00047902 | ✓ |
| | 34. Cingulum_Mid_R | 3.416 | 0.000635417 | ✓ |
| | 63. SupraMarginal_L | 3.391 | 0.000697322 | ✓ |
| | 88. Temporal_Pole_Mid_R | 3.365 | 0.000764793 | ✓ |
| | 19. Supp_Motor_Area_L | 3.314 | 0.000918282 | ✓ |
| | 29, Insula_L | 3.314 | 0.000918282 | ✓ |
| | 18. Rolandic_Oper_R | 3.264 | 0.001099908 | ✓ |
| | 30. Insula_R | 3.238 | 0.001202687 | ✓ |
| | 55. Fusiform_L | 3.238 | 0.001202687 | ✓ |
| | 8. Frontal_Mid_R | 3.213 | 0.001314276 | ✓ |
| | 10. Frontal_Mid_Orb_R | 3.213 | 0.001314276 | ✓ |
| | 52. Occipital_Mid_R | 3.187 | 0.001435352 | ✓ |
| | 33. Cingulum_Mid_L | 3.162 | 0.001566636 | ✓ |
| | 45. Cuneus_L | 3.162 | 0.001566636 | ✓ |
| | 1. Precentral_L | 3.137 | 0.001708899 | ✓ |
| | 50. Occipital_Sup_R | 3.137 | 0.001708899 | ✓ |

| | | | | |
|---|---|---|---|---|
| | 46. Cuneus_R | 3.111 | 0.001862957 | ✓ |
| | 68. Precuneus_R | 3.111 | 0.001862957 | ✓ |
| | 85. Temporal_Mid_L | 3.111 | 0.001862957 | ✓ |
| | 23. Frontal_Sup_Medial_L | 3.086 | 0.002029683 | ✓ |
| | 54. Occipital_Inf_R | 3.060 | 0.00221 | ✓ |
| | 17. Rolandic_Oper_L | 3.035 | 0.002404892 | ✓ |
| | 24. Frontal_Sup_Medial_R | 3.035 | 0.002404892 | ✓ |
| | 41. Amygdala_L | 3.035 | 0.002404892 | ✓ |
| | 49. Occipital_Sup_L | 3.035 | 0.002404892 | ✓ |
| | 69. Paracentral_Lobule_L | 3.035 | 0.002404892 | ✓ |
| | 56. Fusiform_R | 2.984 | 0.002842629 | ✓ |
| | 4. Frontal_Sup_R | 2.933 | 0.003352 | ✓ |
| | 6. Frontal_Sup_Orb_R | 2.933 | 0.003352 | ✓ |
| | 47. Lingual_L | 2.933 | 0.003352 | ✓ |
| | 48. Lingual_R | 2.933 | 0.003352 | ✓ |
| | 57. Postcentral_L | 2.933 | 0.003352 | ✓ |
| | 3. Frontal_Sup_L | 2.908 | 0.003636688 | ✓ |
| | 60. Parietal_Sup_R | 2.908 | 0.003636688 | ✓ |
| | 61. Parietal_Inf_L | 2.908 | 0.003636688 | ✓ |
| | 25. Frontal_Med_Orb_L | 2.857 | 0.004272989 | ✓ |
| | 51. Occipital_Mid_L | 2.857 | 0.004272989 | ✓ |
| | 26. Frontal_Med_Orb_R | 2.832 | 0.004627601 | ✓ |
| | 59. Parietal_Sup_L | 2.806 | 0.005008657 | ✓ |
| | 42. Amygdala_R | 2.781 | 0.005417863 | ✓ |
| | 44. Calcarine_R | 2.730 | 0.006328006 | ✓ |
| | 79. Heschl_L | 2.730 | 0.006328006 | ✓ |
| | 67. Precuneus_L | 2.705 | 0.006832813 | ✓ |
| | 43. Calcarine_L | 2.679 | 0.007373515 | ✓ |
| | 53. Occipital_Inf_L | 2.679 | 0.007373515 | ✓ |
| | 82. Temporal_Sup_R | 2.679 | 0.007373515 | ✓ |
| | 21. Olfactory_L | 2.578 | 0.009940429 | ✓ |
| | 62. Parietal_Inf_R | 2.578 | 0.009940429 | ✓ |
| | 32. Cingulum_Ant_R | 2.527 | 0.011500916 | ✓ |
| | 7. Frontal_Mid_L | 2.502 | 0.012359834 | ✓ |
| | 27. Rectus_L | 2.476 | 0.013275094 | ✓ |
| | 38. Hippocampus_R | 2.476 | 0.013275094 | ✓ |
| | 39. ParaHippocampal_L | 2.476 | 0.013275094 | ✓ |
| | 70. Paracentral_Lobule_R | 2.476 | 0.013275094 | ✓ |
| | 40. ParaHippocampal_R | 2.400 | 0.016390197 | ✓ |
| | 81. Temporal_Sup_L | 2.375 | 0.017562701 | ✓ |
| | 75. Pallidum_L | 2.349 | 0.018808094 | ✓ |
| | 37. Hippocampus_L | 2.324 | 0.020130054 | ✓ |
| | 22. Olfactory_R | 2.299 | 0.021532385 | ✓ |
| | 28. Rectus_R | 2.299 | 0.021532385 | ✓ |
| | 31. Cingulum_Ant_L | 2.197 | 0.028025801 | ✓ |
| | 78. Thalamus_R | 2.070 | 0.038458418 | ✓ |
| **4** | 72. Caudate_R | -3.492 | 0.00047902 | ✓ |
| | 74. Putamen_R | -3.213 | 0.001314276 | ✓ |

| | | | | |
|---|---|---|---|---|
| | 73. Putamen_L | -3.187 | 0.001435352 | ✓ |
| | 56. Fusiform_R | -3.035 | 0.002404892 | ✓ |
| | 23. Frontal_Sup_Medial_L | -2.984 | 0.002842629 | ✓ |
| | 75. Pallidum_L | -2.984 | 0.002842629 | ✓ |
| | 88. Temporal_Pole_Mid_R | -2.984 | 0.002842629 | ✓ |
| | 76. Pallidum_R | -2.857 | 0.004272989 | ✓ |
| | 7. Frontal_Mid_L | -2.832 | 0.004627601 | ✓ |
| | 8. Frontal_Mid_R | -2.806 | 0.005008657 | ✓ |
| | 80. Heschl_R | -2.806 | 0.005008657 | ✓ |
| | 90. Temporal_Inf_R | -2.806 | 0.005008657 | ✓ |
| | 19. Supp_Motor_Area_L | -2.756 | 0.005857017 | ✓ |
| | 85. Temporal_Mid_L | -2.730 | 0.006328006 | ✓ |
| | 71. Caudate_L | -2.629 | 0.00857143 | ✓ |
| | 83. Temporal_Pole_Sup_L | -2.629 | 0.00857143 | ✓ |
| | 29, Insula_L | -2.578 | 0.009940429 | ✓ |
| | 35. Cingulum_Post_L | -2.578 | 0.009940429 | ✓ |
| | 3. Frontal_Sup_L | -2.552 | 0.010695392 | ✓ |
| | 24. Frontal_Sup_Medial_R | -2.552 | 0.010695392 | ✓ |
| | 4. Frontal_Sup_R | -2.476 | 0.013275094 | x |
| | 11. Frontal_Inf_Oper_L | -2.476 | 0.013275094 | X |
| | 34. Cingulum_Mid_R | -2.476 | 0.013275094 | x |
| | 77. Thalamus_L | -2.451 | 0.014249763 | x |
| | 16. Frontal_Inf_Orb_R | -2.426 | 0.015287028 | x |
| | 87. Temporal_Pole_Mid_L | -2.426 | 0.015287028 | x |
| | 54. Occipital_Inf_R | -2.400 | 0.016390197 | x |
| | 14. Frontal_Inf_Tri_R | -2.375 | 0.017562701 | x |
| | 13. Frontal_Inf_Tri_L | -2.349 | 0.018808094 | x |
| | 86. Temporal_Mid_R | -2.324 | 0.020130054 | x |
| | 30. Insula_R | -2.299 | 0.021532385 | x |
| | 10. Frontal_Mid_Orb_R | -2.248 | 0.024593987 | x |
| | 15. Frontal_Inf_Orb_L | -2.222 | 0.026261484 | x |
| | 65. Angular_L | -2.222 | 0.026261484 | x |
| | 61. Parietal_Inf_L | -2.197 | 0.028025801 | x |
| | 78. Thalamus_R | -2.197 | 0.028025801 | x |
| | 36. Cingulum_Post_R | -2.121 | 0.033944438 | x |
| | 81. Temporal_Sup_L | -2.095 | 0.03614139 | x |
| | 55. Fusiform_L | -2.070 | 0.038458418 | x |
| | 89. Temporal_Inf_L | -2.019 | 0.043472754 | x |
| | 12. Frontal_Inf_Oper_R | -1.994 | 0.046180343 | x |
| | 9. Frontal_Mid_Orb_L | -1.968 | 0.049028558 | x |
| | 82. Temporal_Sup_R | -1.968 | 0.049028558 | x |
| **5** | 1. Precentral_L | 2.756 | 0.005857017 | x |
| | 86. Temporal_Mid_R | 2.679 | 0.007373515 | x |
| | 2. Precentral_R | 2.476 | 0.013275094 | x |
| | 20. Supp_Motor_Area_R | 2.476 | 0.013275094 | x |
| | 37. Hippocampus_L | 2.451 | 0.014249763 | x |
| | 88. Temporal_Pole_Mid_R | 2.222 | 0.026261484 | x |
| | 58. Postcentral_R | 2.197 | 0.028025801 | x |

| | | | | |
|---|---|---|---|---|
| | 64. SupraMarginal_R | 2.121 | 0.033944438 | x |
| | 3. Frontal_Sup_L | 2.070 | 0.038458418 | x |
| | 41. Amygdala_L | 2.045 | 0.040900509 | x |
| | 59. Parietal_Sup_L | 2.019 | 0.043472754 | x |
| | 82. Temporal_Sup_R | 2.019 | 0.043472754 | x |
| | 18. Rolandic_Oper_R | 1.994 | 0.046180343 | x |
| | 38. Hippocampus_R | 1.968 | 0.049028558 | x |
| **6** | - | | | |
| **7** | 87. Temporal_Pole_Mid_L | 2.222 | 0.026261484 | x |
| | 34. Cingulum_Mid_R | -2.070 | 0.038458418 | x |

***Supplementary Table S4. Rumination MBCT+TAU differences in projection of brain dynamics into each manifold network.*** *We computed the difference in projection maps between before and after treatment. The significant contributions for each latent are shown in each row, identified as brain area, change (positive for stronger values after treatment), p-value and whether they survive to correction by multiple comparisons.*
